# Atrial Fibrillation as a Determinant of Brain Health: Multimodal Evidence Supports Stroke-Dependent and Stroke-Independent Effects

**DOI:** 10.64898/2026.08.11.26360168

**Authors:** Alison Offer, Parag R. Gajendragadkar, Cornelia Van Duijn, Paul M. Matthews, Barbara Casadei, Jemma C Hopewell

**Affiliations:** Clinical Trial Service Unit and Epidemiological Studies Unit, Nuffield Department of Population Health, University of Oxford, Oxford, United Kingdom; UK Dementia Research Institute Centre, Department of Brain Sciences, Imperial College London and Rosalind Franklin Institute, Rutherford Appleton Laboratory, Didcot, UK; National Heart and Lung Institute, Imperial College London, London, United Kingdom

**Keywords:** Epidemiology, Atrial Fibrillation, Brain imaging, Cognition, Dementia, Stroke

## Abstract

**Background and Aims:** Atrial fibrillation (AF) is associated with dementia and cognitive decline; it remains unclear whether the association is independent of cardiovascular comorbidities, particularly stroke. We aimed to establish stroke-dependent and stroke-independent effects of AF on multiple layers of brain health.

**Methods:** We investigated stroke-dependent and stroke-independent associations of AF with brain MRI imaging measures, cognitive performance, incident dementia, and biomarkers of neuronal and glial injury among 502 099 participants in the UK Biobank. Genetic analyses were performed to support causal inference.

**Results:** AF was associated with lower global and regional grey matter volume. After full adjustment, AF remained associated with lower grey matter volume (-0.13 SD, 95%CI:-0.15 to-0.10), greater white matter hyperintensity burden (0.06 SD, 0.03 to 0.10), and higher mean diffusivity (0.06 SD, 0.03 to 0.10). Genetic analyses were consistent with stroke-independent associations between AF and grey matter loss. AF was associated with poorer cognitive performance and higher concentrations of neurofilament light chain, a marker of neuronal injury. Associations with vascular and all-cause dementia were attenuated by adjustment for cardiovascular risk-factors and stroke. Genetic analyses supported ischaemic stroke as a major driver of dementia risk in AF.

**Conclusion:** Associations with grey matter and cognitive impairment persisted after accounting for stroke, whereas the association with dementia was largely explained by ischaemic stroke. This highlights the potential for AF to impact brain health through both stroke-dependent and stroke-independent mechanisms; stroke prevention is necessary but may not be sufficient to fully address the broader burden of AF-associated brain injury.

## Introduction

Atrial fibrillation (AF) is the most common sustained cardiac arrhythmia, affecting an estimated 33 million people globally^1,2^ with a prevalence of ∼4% in Europeans aged 60–70 and 10–17% in those over 80.^3^ AF is a major risk factor for cardioembolic stroke, which is associated with accelerated cognitive decline and higher dementia risk.^4–7^ Although anticoagulation therapy reduces the risk of overt cardioembolic stroke, individuals with AF continue to experience silent cerebral infarcts and cognitive deterioration despite anticoagulation.^8,9^ The BRAIN-AF trial examining the impact of low dose rivaroxaban on neurocognitive impairment in AF patients at low stroke risk was stopped early due to futility.^10^ Notably, a significant association between AF and all-cause dementia appears to persist, even after accounting for prior stroke.^11–25^

The mechanism linking AF to impaired brain health is likely multifactorial. Individuals with AF frequently present with atherosclerosis, heart failure, and other cardiovascular risk factors that may contribute to lacunar infarcts, small vessel disease, and chronic cerebral hypoperfusion, which accelerate cognitive decline.^4^ Neuroimaging studies have provided insights into associated pathology, demonstrating associations between AF and cortical atrophy, microbleeds, and imaging markers of small vessel disease and neurodegeneration, including white matter hyperintensities (WMH),^26–32^ while recent advances in proteomic profiling offer opportunities to detect subclinical brain injury.^33^

We have leveraged the large-scale multimodal data available in the UK Biobank (UKB)^34^ to (1) determine the relationship between AF and measures of brain health, (2) the extent to which cardiovascular comorbidities explain these, and (3) identify potential strategies to mitigate the cognitive burden associated with AF in a population setting.

## Methods

### Participants

The UK Biobank (UKB) is a population-based prospective cohort of ∼500 000 individuals who were recruited between 2006 and 2010^34^ with available follow-up to 2023. Baseline data included lifestyle, medical history, cognition, physical measures, and genetics. Linked data includes neuroimaging, proteomics, death records, and hospital admissions, diagnoses, and procedures (HADP) data. This analysis considers 502 099 participants with no record of pre-recruitment dementia. Sub-cohorts were defined for brain imaging, cognitive function, and dementia (**eFigure 1 in the Supplement**).

### AF events in UKB

Atrial fibrillation was defined by the first self-reported AF or flutter, or first AF-related ICD-10/OPCS-4 code in the HADP data, comprising English Hospital Episode Statistics (1997–31 March 2023), Welsh Patient Episode Data (1992–31 May 2022), and Scottish Morbidity Records (1989–31 Aug 2022) (**eTable 1 in the Supplement**).

### Outcomes

Primary outcomes were (i) global and regional brain imaging markers from structural-weighted and diffusion-weighted brain MRI, (ii) cognitive function measured by online cognitive function tests, and (iii) incident dementia.

*(i)* *Brain Imaging*

The imaging cohort comprised UKB participants with pre-processed structural T1-weighted MRI, T2-weighted fluid-attenuated inversion recovery (FLAIR), and diffusion MRI data.^35,36^ Volumetric imaging-derived phenotypes (IDPs) included global brain volume parameters (total brain volume, total and peripheral cortical grey matter, total white matter, total cerebrospinal fluid (CSF)), grey matter volumes in cortical structures and the cerebellum, (with tissue segmentation performed using FAST ^37^), subcortical volumes (segmented using FIRST ^38^), and log-transformed WMH volume (estimated using BIANCA^39^). These were normalised for head size prior to analysis. The diffusion MRI-derived markers comprised global fractional anisotropy (gFA) and global mean diffusivity (gMD),^40^ obtained via factor analysis across 27 white matter tracts (**eMethods in the Supplement**). Higher gMD and lower gFA can indicate reduced white matter integrity related to axonal or myelin degradation. As of November 2025, IDPs were available for ∼83,000 participants. After technical exclusions (**eMethods**), the imaging cohort included 72 973 individuals, with 2538 AF cases before the imaging visit (**eFigure 1**).

*(ii)* *Cognitive Function*

Cognitive assessment was conducted during the imaging visits and via online assessments in 2014/5 and 2021/2. Here, data from the second online assessment was analysed to maximise follow-up and case accrual. Tests included fluid intelligence (FI), digit symbol substitution (DSS), matrix pattern completion (MPC), and trail making (TM; log-transformed times).^41^ The cognitive cohort comprised 180 660 UKB participants who completed at least one online test in 2021-2022; 9606 had an AF diagnosis before the cognitive assessment (**eFigure 1**).

*(iii)* *Dementia*

Dementia was defined as the first dementia-related ICD-10 code in routine hospital admission data, death records, or self-report and was subclassified through ICD-10 codes, as Alzheimer’s disease, vascular, frontotemporal or undefined/mixed dementia (**eTable 2 in the Supplement**). When two subtypes were recorded, cases were classified as “mixed” if both appeared at the first record; otherwise, classification followed the first available code. Dementia has a long prodromal phase and the precise onset dates and ordering of both HADP-sourced AF and dementia are uncertain, therefore the analyses considered AF recorded before the age of 65 as the exposure and the first dementia diagnosis after age 65 as the outcome. The dementia cohort comprised 365 979 participants who were older than 65 at the end of follow-up and free of dementia before turning 65 (**eFigure 1**); 10 462 of these had AF reported before age 65.

### Protein markers of neuronal damage

UKB measured neurofilament light chain (NfL) and glial fibrillary acidic protein (GFAP), which have been associated with neuronal damage and incident dementia,^42–44^ in around 50,000 individuals at recruitment using the Olink Target 36 platform. After excluding non-randomly selected participants, plasma NfL and GFAP measurements were available for approximately 44,300 individuals, including 732 with pre-recruitment AF and 242 with pre-recruitment stroke (**eMethods**).

This study was reported in accordance with the STROBE^45^ guidelines for observational studies.

### Statistical methods

Linear regression was used to analyse the associations between diagnosis of AF before the imaging visit and the brain imaging IDPs, and between diagnosis of AF before cognitive assessment and the cognitive scores. Cox regression was used to investigate the association between a first report of AF before age 65 and a first report of dementia after age 65 (**eMethods**). Two models were fitted for each outcome: The first adjusted for demographics (age, sex, Townsend deprivation index, education) and, where relevant, technical covariates (e.g., imaging-related covariates), the second further adjusted for cardiovascular risk factors (smoking, alcohol, body mass index (BMI), systolic blood pressure, LDL-, HDL-, and total cholesterol, any diabetes) and cardiovascular comorbidities (heart failure (HF), coronary heart disease (CHD), haemorrhagic and (separately) ischaemic stroke). Covariate details are summarised in **eTable 3 in the Supplement**; definitions of the stroke, HF, CHD and diabetes phenotypes are given in **eTable 4 and eTable5 in the Supplement**. P-value thresholds for significance were adjusted for multiple testing within each analysis cohort and reported in the figure captions (**eMethods**).

### Genetic analysis methods

Secondary analyses based on genetic data were performed using an inverse-variance weighted (random effects) two-sample Mendelian randomization (MR) framework. Genetically proxied AF was derived from genome-wide significant variants from a recent AF meta-analysis,^46^ clumped, and harmonised with UKB genetic variants (**eMethods**), yielding 269 genetic variants; outcome data were sourced from UKB participants as described above. To determine effects independent of the effects on ischaemic stroke, estimates were also adjusted for the effects of the same 269 genetic variants on total ischaemic stroke, based on summary statistics from the *GIGASTROKE* Consortium,^47^

## Results

The imaging and cognitive cohorts (**eFigure 1, Table 1**) were younger than the overall UKB population (mean age at recruitment 54.5 and 55.4 years, respectively, versus 56.5 years; **Table 1**), came from less deprived areas and were more often degree-educated. Conversely, the dementia cohort, by design, was older (restricted to participants aged 65 or over by the end of the HADP-linked follow-up (2023) and otherwise closely matched the overall UKB population.

**Table 1:** Characteristics at the UKB recruitment visit, overall and by analysis cohort. ^a^ After removing individuals with pre-recruitment dementia. ^b^ Individuals with records of AF before the imaging visit, cognitive assessment, or before 65th birthday respectively. ^c^ Missingness < 0.3%. ^d^ Missingness < 0.6%

|  | All UKB<sup>a</sup><br>(N=502099) | Brain imaging cohort<br>(N=72973) | Cognitive cohort<br>(N=180660) | Dementia cohort<br>(N=365879) |
| --- | --- | --- | --- | --- |
| Individuals with AF<sup>b</sup>, n(%) |  | 2538 (3.4%) | 9606 (5.3%) | 10462 (2.8%) |
| Baseline Age (years) |  |  |  |  |
| mean (SD) | 56.5 (8.1) | 54.5 (7.5) | 55.4 (7.6) | 60.5 (5.23) |
| <60, n (%) | 284815 (56.7%) | 50746 (69.7%) | 117157 (64.8%) |  |
| 60+, n (%) | 217284 (43.3%) | 22042 (30.3%) | 63503 (35.2%) |  |
| Age at imaging, mean (SD) |  | 66.08 (7.9) |  |  |
| Female sex, n (%) | 273176 (54.4%) | 39208 (53.9%) | 103183 (57.1%) | 198896 (54.4%) |
| Smoking status, n (%) |  |  |  |  |
| Current | 52935 (10.5%) | 4535 (6.2%) | 13419 (7.4%) | 33465 (9.1%) |
| Former | 172899 (34.4%) | 23793 (32.7%) | 62861 (34.8%) | 138650 (37.9%) |
| Never | 273318 (54.4%) | 44291 (60.8%) | 103920 (57.5%) | 191568 (52.4%) |
| Unknown | 2947 (0.6%) | 169 (0.2%) | 460 (0.3%) | 2196 (0.6%) |
| Alcohol Consumption, n (%) |  |  |  |  |
| Under 3 units/week | 95072 (18.9%) | 10442 (14.3%) | 29125 (16.1%) | 68913 (18.8%) |
| 3-7 units/week | 101559 (20.2%) | 16896 (23.2%) | 41526 (23.0%) | 75203 (20.6%) |
| 8-15 units/week | 111271 (22.2%) | 19120 (26.3%) | 45590 (25.2%) | 81382 (22.2%) |
| 16+ units/week | 112193 (22.3%) | 16713 (23.0%) | 40128 (22.2%) | 81649 (22.3%) |
| Unknown | 82004 (16.3%) | 9617 (13.2%) | 24291 (13.4%) | 58732 (16.1%) |
| Systolic Blood pressure (mmHg)<sup>c</sup> |  |  |  |  |
| Mean (SD) | 137.9 (18.7) | 134.5 (176) | 135.8 (17.9) | 140.8 (18.7) |
| Body mass index (kg/m<sup>2</sup>)<sup>d</sup> |  |  |  |  |
| Mean (SD) | 27.4 (4.8) | 26.4 (4.1) | 26.8 (4.6) | 27.5 (4.7) |
| Education, n (%) |  |  |  |  |
| Degree | 162453 (32.4%) | 33982 (46.7%) | 79155 (43.8%) | 109910 (30.0%) |
| Professional or post-16 | 163383 (32.5%) | 24299 (33.4%) | 61103 (33.8%) | 115539 (31.6%) |
| None of these | 176263 (35.1%) | 14507 (19.9%) | 40402 (22.4%) | 140430 (38.4%) |
| Townsend deprivation index, Mean (SD) | -1.3 (3.1) | -1.9 (2.7) | -1.7 (2.8) | -1.5 (3.0) |

### AF and brain imaging markers

After adjustment for demographic and technical covariates (**eMethods**), a diagnosis of AF before brain MRI was strongly associated (P<.0001) with lower total grey matter (estimate=-0.16SD), peripheral cortical grey matter volumes and higher cerebrospinal fluid volume (**Figure 1**). AF was also associated with greater WMH volume (estimate=0.08SD, P<.0001) and higher global mean diffusivity (estimate=0.09SD, P<0.0001), and lower global fractional anisotropy (estimate=-0.05SD, P=0.001) indicating lower white matter integrity. By contrast, no association was observed with total white matter volume.

**Figure 1:**
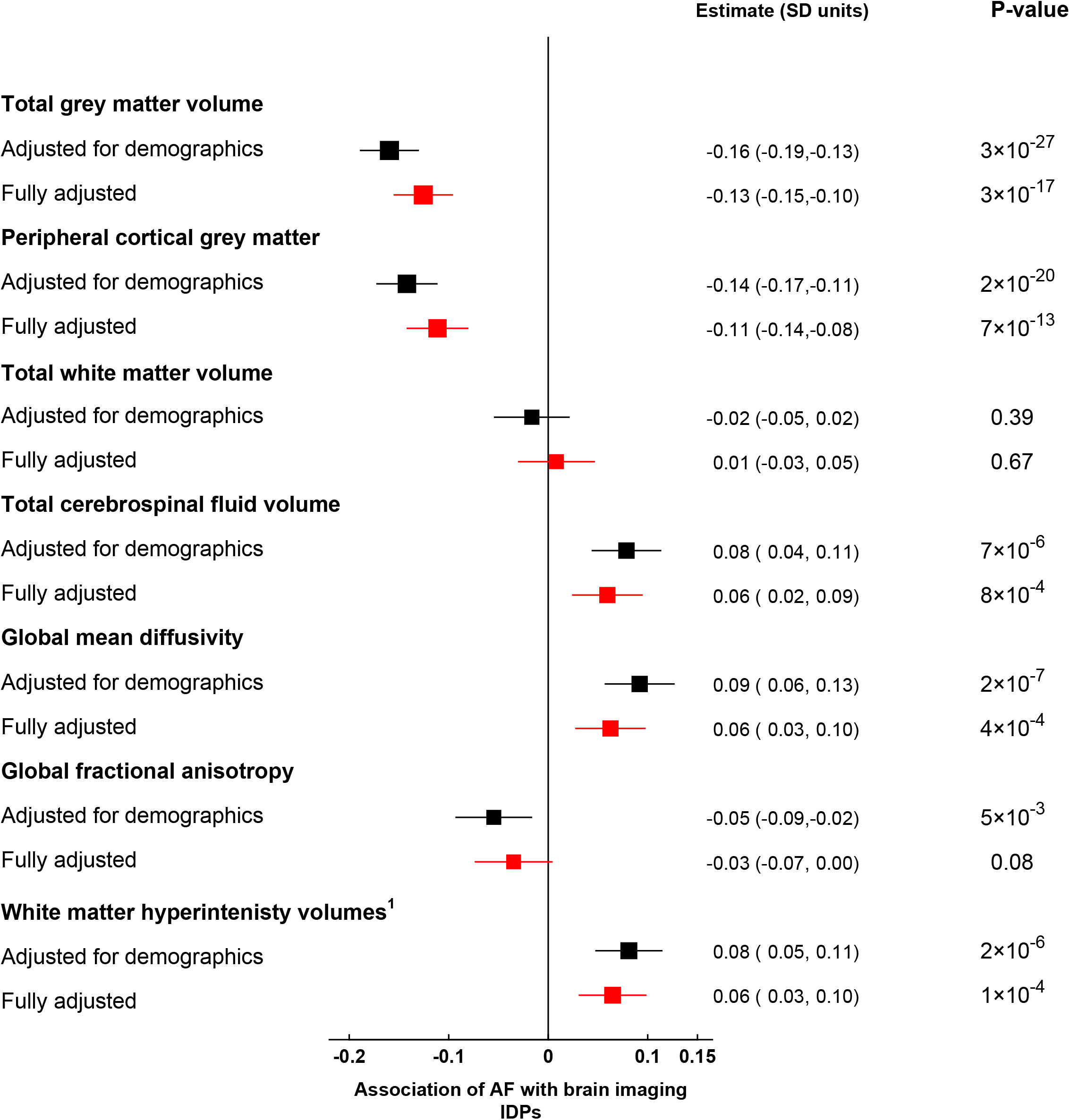
Observational association of AF with global imaging IDPs in 72 973 individuals. ^a^ White matter hyperintensity volumes are log transformed. Associations are with AF events (n=2538) recorded at any time before the imaging visit. Associations were adjusted for age, sex, education, and the Townsend deprivation index at recruitment, plus technical imaging confounders. Full adjustment also included baseline CVD risk factors and other CVD comorbidities diagnosed before the imaging visit, including haemorrhagic and ischaemic stroke. A Bonferroni corrected P-value threshold for significance of 7×10^-3^ is assumed. **Alt Text**:Forest plot showing the associations between AF and total and peripheral grey matter volume, white matter volume, CSF volume, mean diffusivity, fractional anisotropy and WMH volumes, showing results adjusted for demographics and fully adjusted.

Adjustment for cardiovascular risk factors and comorbidities, including prior stroke, attenuated all significant associations by between 18% for total grey matter volume and 40% for global fractional anisotropy. However, the relationships between AF and smaller grey matter volume (estimate=−0.13 SD, P<.0001), higher WMH burden (estimate=0.06 SD, P=.0001), and elevated mean diffusivity (estimate=0.06 SD, P=0.0004) remained highly significant (**Figure 1**), suggesting that AF is independently associated with both cortical atrophy and these imaging markers of small vessel disease.

At a regional level, after adjustment for demographic and technical covariates, AF was consistently associated with lower regional cortical grey matter volumes across the cortical regions (frontal, parietal, temporal and occipital) and the cerebellum (**eFigure 2 in the Supplement**), with reductions of up to 0.17 SD. AF was also associated with smaller subcortical volumes, most notably in the hippocampus and thalamus, while the amygdala was the only structure not associated with AF (**eFigure 2**). Full adjustment reduced the associations, but the pattern remained.

Genetically proxied AF showed an association with total and peripheral grey matter volumes (estimates=−0.02 SD per log-odds of AF, P≤.003) and global mean diffusivity (estimate=0.03, P=0.0001), but the association with log-transformed WMH volume did not reach significance (**Table 2**). After adjusting for the effects of the genetic variants on ischaemic stroke, to account for potential interdependencies with brain measures, the associations with white matter integrity measures were completely attenuated. By contrast, the association with grey matter volume remained significant (−0.03; P<.0001), supporting the relationship between AF and cortical atrophy that is independent of ischaemic stroke.

**Table 2:**
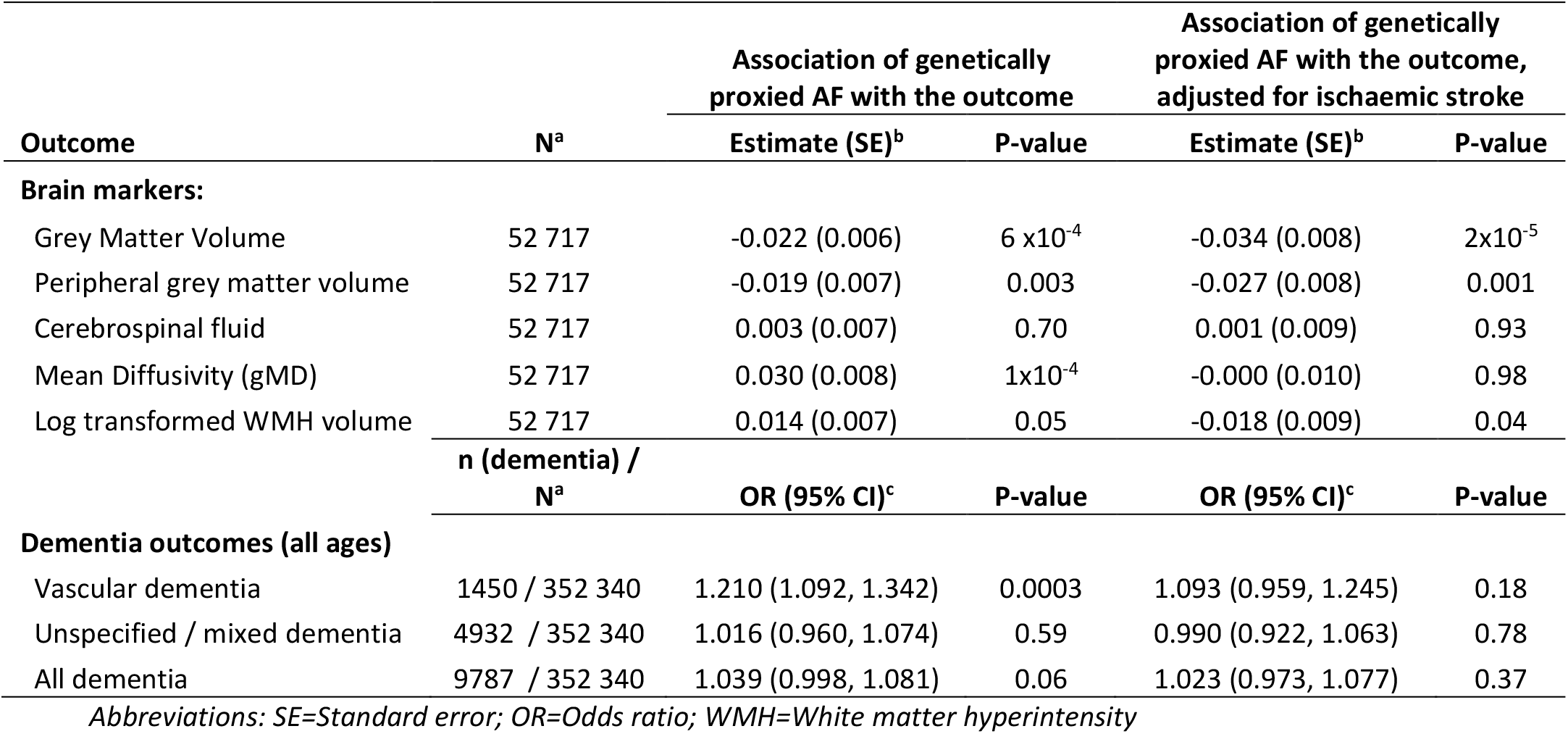
Associations of genetically proxied AF with imaging and dementia outcomes Genetic analyses were restricted to associations that reached statistical significance in the observational analysis. ^a^ Number of European non-related individuals with genetic information and non-missing values of the outcome; ^b^ units of standard deviation of outcome per unit higher in log odds of atrial fibrillation; ^c^ Odds ratio per unit higher in log odds of atrial fibrillation

### AF and cognitive performance

Of the surviving UKB participants invited to undertake the second online cognitive assessment, 180,660, with a mean age of 68.1 years, completed at least one test, including 9606 individuals with a prior diagnosis of AF (**eFigure 1**).

After adjustment for demographics and education, a record of AF before the cognitive assessment was associated with small but significant (P<.0001) reductions in cognitive performance across all four tests (0.05–0.10 SD; **Figure 2**). After additional adjustment for cardiovascular risk factors and comorbidities, including ischaemic stroke, AF remained independently associated with three of the four cognitive test scores (P<.01), although effect sizes were reduced to ≤ 0.05 SD. There was no association between genetically proxied AF and cognitive scores (**eTable 6 in the Supplement**).

**Figure 2:**
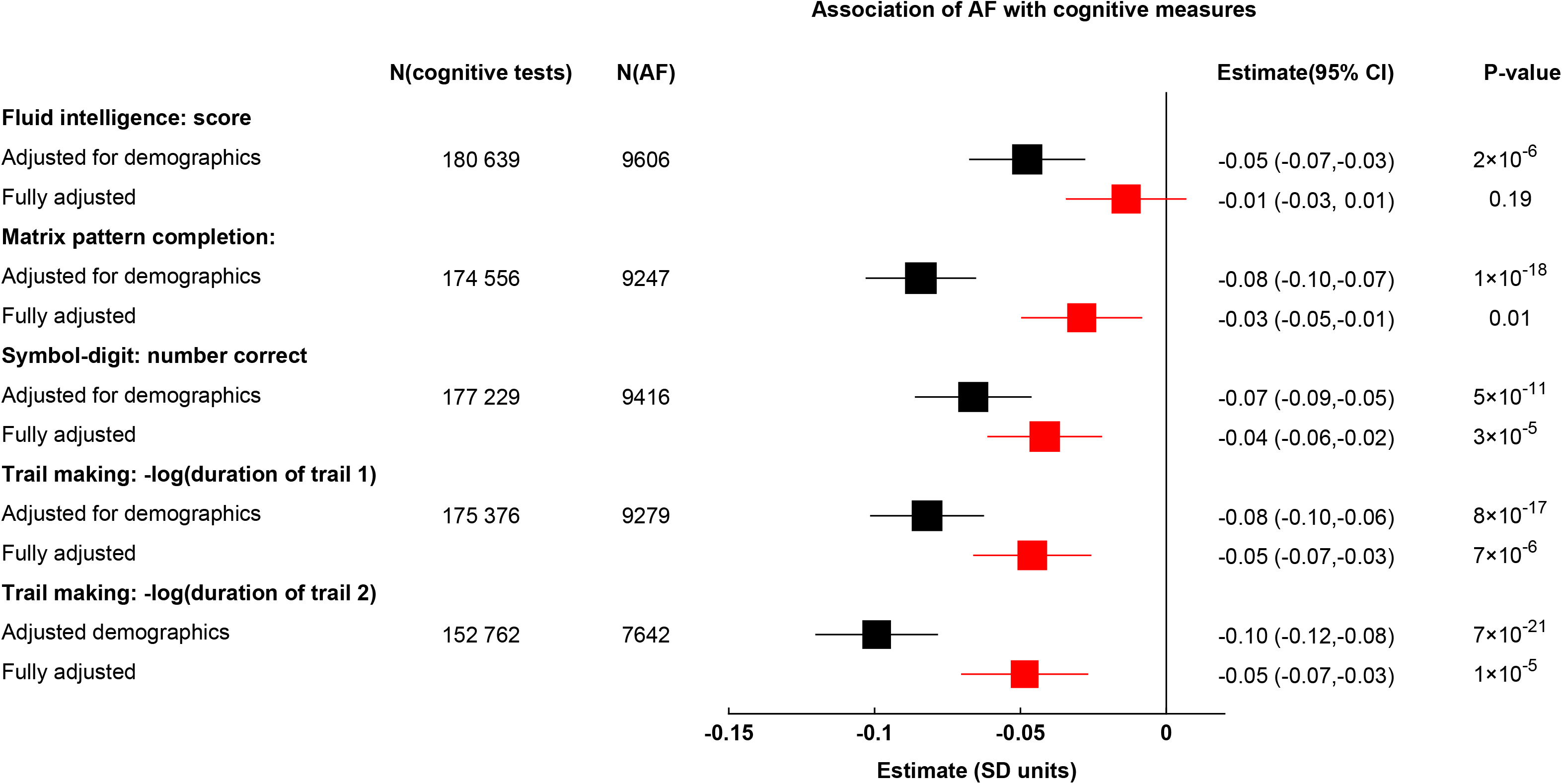
Observational association of AF with cognitive measures. Results are based on AF diagnosed before the cognitive assessment with the second online cognitive assessment in UK Biobank (conducted between 2021 and 2022). Associations were adjusted for age at cognitive assessment, sex, education and recruitment Townsend deprivation index. Full adjustment also included baseline CVD risk factors and other CVD comorbidities diagnosed before the assessment, including haemorrhagic and ischaemic stroke. A Bonferroni corrected P-value threshold for significance of 0.01 is assumed. **Alt Text:** Forest plot showing the associations between AF and individual cognitive test scores, showing results adjusted for demographics and fully adjusted.

### AF and risk of dementia

Among 365 979 participants in the dementia cohort, with a mean follow-up of 7.8 years, 10 462 had AF before age 65 (**eTable 7 in the Supplement**). After adjustment for demographics and education, AF (before age 65) was associated with higher risk of vascular dementia (after age 65) (HR 1.7; 95% CI, 1.4, 2.1; P<.0001). It was also associated with unspecified/mixed and all-cause dementia (**Figure 3**). No association was observed with Alzheimer’s disease. Adjustment for cardiovascular risk factors and comorbidities markedly attenuated the association with vascular dementia (HR 1.2; 95% CI, 1.0, 1.5; P=.10), with similar attenuation for unspecified/mixed and all-cause dementia.

**Figure 3:**
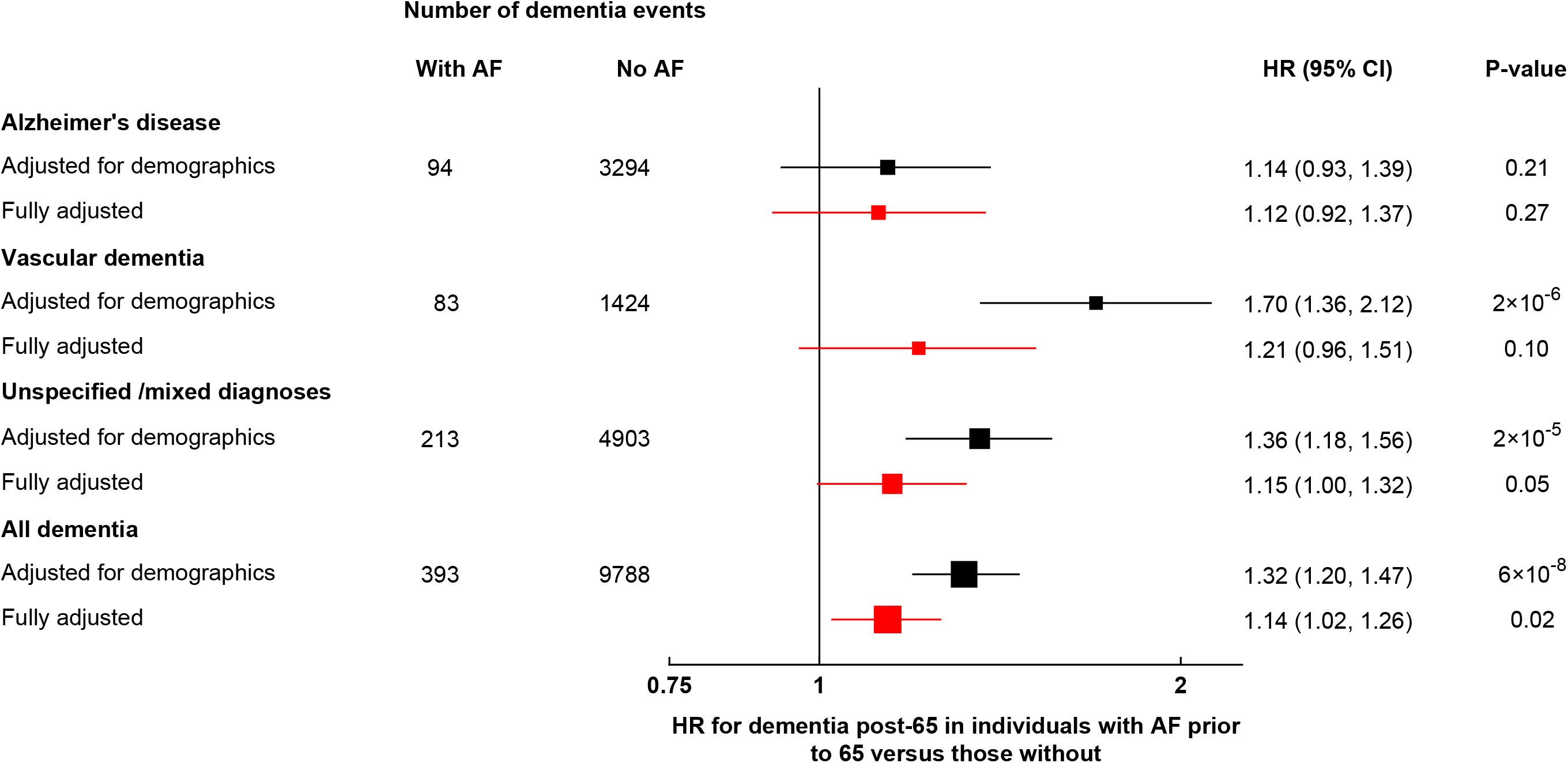
Observational association of AF with dementia among 365 979 individuals. Associations are between first report of dementia after the age of 65 and AF diagnosed before the age of 65. Associations were analysed using age-at-risk as the timescale, also adjusted for year of recruitment, sex Full adjustment also included CVD risk factors at recruitment, prevalent and incident diabetes mellitus, heart failure, CHD, haemorrhagic stroke, and ischaemic stroke.The small number of frontal temporal dementia events (n=176, three in participants with a prior diagnosis of AF) is not shown separately, though they were included in the “all dementia” results., mean number of hospitalisations, education, and the Townsend deprivation index. A Bonferroni adjusted P-value for significance of 0.0125 is assumed. **Alt Text**:Forest plot showing the associations between AF and Alzheimer’s disease, vascular, unspecified/mixed, and all dementia, showing results adjusted for demographics and fully adjusted.

Genetic analyses showed a significant association only between genetically proxied AF and vascular dementia events (OR 1.21 per log-odds higher risk of AF; 95% CI, 1.10, 1.34; P=.0002, **Table 2**). This association was attenuated by about half and no longer significant after adjusting for the genetic effects of ischaemic stroke (OR 1.09; 95% CI, 0.96, 1.25; P=.18), although the confidence interval does not exclude a residual role of AF in dementia risk independent of ischaemic stroke.

### AF and protein markers of neuronal damage

NfL is a non-specific marker of neuronal damage,^48^ and has been previously associated with AF in a small stroke-free cohort,^49^ whereas GFAP, a marker of astroglial injury that is elevated in the presence of stroke, is an independent predictor of future dementia.^42,43^ In our cohort, prevalent AF was associated with NfL levels (0.09 NPX units; 95%CI, 0.06, 0.13; P<.0001), equivalent to a 6.7% (4.2%, 9.3%) higher NfL level (**eMethods**), independent of ischaemic stroke and other cardiovascular comorbidities (**eFigure 3 in the Supplement**). By contrast, AF showed no association with GFAP levels (–0.01 NPX units; 95% CI, –0.04, 0.03; P=.56), despite a strong association of GFAP with prevalent ischaemic stroke (0.18 NPX units; 95% CI, 0.10, 0.25; P<.0001), equivalent to a 13.0% higher GFAP level (**eMethods**). Results were unaltered by further adjustment for estimated glomerular filtration rate, which can affect these markers.

## Discussion

In this large, multimodal analysis of UK Biobank participants, AF is associated with structural brain changes, modest cognitive deficits, and a higher risk of vascular dementia. By integrating observational analyses, genetic analyses, and proteomic biomarkers, we were able to distinguish stroke-dependent and stroke-independent contributions in relation to AF and adverse brain outcomes. Overall, AF is robustly associated with widespread grey matter atrophy, higher NfL, and a modest reduction in cognitive performance - all of which persist after full adjustment for cardiovascular comorbidities, including prior stroke. The regional pattern—affecting all major cortical lobes, the cerebellum, and key subcortical structures such as the hippocampus and thalamus—indicates widespread vulnerability of grey matter in individuals with AF. The observational findings also suggest a residual association between AF and brain tissue health (as measured by global mean diffusivity) and the total volume of WMH after adjustment for cardiovascular comorbidities, although genetic analyses did not support a causal interpretation. Ischaemic stroke and other cardiovascular comorbidities explained much of the association between AF and higher dementia risk.

Our large-scale findings are supported by more limited studies in specific patient groups suggesting associations between AF and smaller localised brain volumes in stroke-free patients.^26,28,30,32,50,51^ A Mendelian randomization study, using earlier and notably smaller UKB datasets and weaker genetic instruments for AF, concluded that genetic predisposition to AF was associated with lower total grey matter (but not white matter) volume and suggested this may be mediated by ischaemic stroke.^43^ A recent study suggested that non-lacunar infarcts, periventricular WMH, and fewer cerebral microbleeds may differentiate AF-associated brain changes from those that are more atherosclerotic in nature.^8^ These findings are consistent with the idea that AF alone can lead to specific imaging changes.

Previous studies have found associations between AF, cognition, and dementia.^6,52–54^ However, we have found for the first time that AF is associated with effects across multiple cognitive domains. Effect sizes were modest and attenuated after adjustment for vascular comorbidities, but the persistence of associations for most tests suggests that AF may contribute to subtle cognitive decline before the onset of clinical dementia. Plasma biomarker analyses provided additional mechanistic insights. AF was associated with higher plasma NfL, a marker of neuronal injury, independent of stroke and other comorbidities. By contrast, AF was not associated with GFAP. This suggests that AF may contribute to neuronal injury through non-stroke mechanisms, while glial activation, and much of the dementia risk, appears to be driven by cerebrovascular events, though other factors, related to the time course of pathology and clearance of the markers, may be important.

A clear understanding of the impact of AF on brain health has been hampered by varying study designs in the existing literature. For example, in recent reports using subsets of the UKB data, Zhai^55^ and Ou^56^ found significant associations between a diagnosis of AF preceding enrolment in UKB and all-cause, Alzheimer’s and vascular dementia on follow-up; however, they either did not consider the role of stroke^56^ or failed to fully adjust, by only considering stroke prior to recruitment.^55^ Our findings are consistent with those of an earlier Mendelian randomisation study,^57^ which suggested a 4% (95% CI, 2%, 27%) higher risk of all-cause dementia per log-odds-higher genetically proxied AF risk, which attenuated to a non-significant 7% (95% CI, −5%, 20%) effect after adjustment for ischaemic stroke or low cardiac output. Genetically proxied AF was not associated with cognitive performance in the present study, indicating that the cognitive effects observed in mid-to-late life may be mediated by comorbidities, cumulative vascular injury, or the duration and severity of AF rather than AF liability itself. Studies have shown that AF is associated with adverse changes in cerebral blood flow, which reverse upon restoration of sinus rhythm.^58,59^ Whether these changes are sufficient to explain residual associations between AF, grey matter atrophy, and cognitive function merits further investigation. Similarly, whilst it has been established that anticoagulation reduces the risk of clinical stroke associated with AF, some studies suggest that restoration of sinus rhythm on top of anticoagulation may be associated with small incremental benefits in reducing cerebral events.^52^

This study leverages a large UK Biobank cohort with multimodal phenotyping, in a predominantly European population, which may limit generalisability. Exposure and outcome capture also have limitations: HADP lacks detailed AF subtype classification and often lags disease onset, especially for outcomes such as AF and dementia, which do not typically present initially in secondary care. The AF cases among the younger, healthier sub-cohort with imaging data may be predominantly paroxysmal, low-burden AF, which could understate associations with brain imaging markers.^60^ Follow-up in the dementia cohort may still be too short for all dementias to develop beyond the prodromal phase, and the absence of comprehensive prescribing data precludes assessment of anticoagulation effects at the current time.

The UK Biobank is currently requesting that individuals over the age of 65 who attend imaging visits wear a heart monitor for two weeks to detect subclinical arrhythmias. These data will provide important new opportunities to investigate the associations between AF burden, brain imaging, and cognitive function, alongside comprehensive prescribing data that will emerge when complete primary care data are made available that will also allow assessment of anticoagulation effects that are not currently possible.

Our population-based findings support a model where stroke prevention remains central to reducing the risk of AF-related adverse brain outcomes, but may not fully address the broader burden of AF-associated brain injury. In this paradigm, the persistence of associations with grey matter loss, cognitive performance, and neurofilament light chain after adjustment for stroke and cardiovascular comorbidities supports evaluation of additional AF-directed strategies, including earlier detection, improved risk-factor control, and rhythm-or burden-reduction approaches, for preservation of brain health. Thus, AF management requires a brain-health framework beyond overt stroke prevention, that will mitigate its broader impact on brain health.

## Acknowledgments and Funding

This research was conducted using the UK Biobank Resource under application number 14568. The authors acknowledge support from the National Institute for Health and Care Research (NIHR) Oxford Biomedical Research Centre (BRC), British Heart Foundation, and Nuffield Department of Population Health, University of Oxford, UK.

## Data availability

Data used in this study was derived from the UK biobank resource under approved application number 14568. UK Biobank data are available to bona fide researchers via application to UK Biobank.

## References

1. Chugh SS, Havmoeller R, Narayanan K, et al. Worldwide epidemiology of atrial fibrillation: a Global Burden of Disease 2010 Study. Circulation 2014; 129(8): 837–47.

2. Chung MK, Eckhardt LL, Chen LY, et al. Lifestyle and Risk Factor Modification for Reduction of Atrial Fibrillation: A Scientific Statement From the American Heart Association. Circulation 2020; 141(16): e750–e72.

3. Zoni-Berisso M, Lercari F, Carazza T, Domenicucci S. Epidemiology of atrial fibrillation: European perspective. Clin Epidemiol 2014; 6: 213–20.

4. Rivard L, Friberg L, Conen D, et al. Atrial Fibrillation and Dementia: A Report From the AF-SCREEN International Collaboration. Circulation 2022; 145(5): 392–409.

5. Bunch TJ. Atrial Fibrillation and Dementia. Circulation 2020; 142(7): 618–20.

6. Alonso A, Arenas de Larriva AP. Atrial Fibrillation, Cognitive Decline And Dementia. Eur Cardiol 2016; 11(1): 49–53.

7. Dietzel J, Haeusler KG, Endres M. Does atrial fibrillation cause cognitive decline and dementia? Europace 2018; 20(3): 408–19.

8. Stegmann T, Joundi RA, Srivastava A, et al. Atrial fibrillation and atherosclerosis cause different vascular brain lesions on magnetic resonance imaging. Eur Heart J 2025; 46(47): 5177–88.

9. Svennberg E, Merino JL, Andrade J, et al. Transforming atrial fibrillation management by targeting comorbidities and reducing atrial fibrillation burden: the 10th AFNET/EHRA consensus conference. Europace 2025; 27(12).

10. Rivard L, Khairy P, Talajic M, et al. Anticoagulation to prevent ischemic stroke and neurocognitive impairment in atrial fibrillation: the BRAIN-AF randomized clinical trial. Nat Med 2026; 32(1): 297–305.

11. Dublin S, Anderson ML, Haneuse SJ, et al. Atrial Fibrillation and Risk of Dementia: A Prospective Cohort Study. Journal of the American Geriatrics Society 2011; 59(8): 1369–75.

12. Zhang W, Liang J, Li C, et al. Age at Diagnosis of Atrial Fibrillation and Incident Dementia. JAMA Netw Open 2023; 6(11): e2342744.

13. Giannone ME, Filippini T, Whelton PK, et al. Atrial Fibrillation and the Risk of Early-Onset Dementia: A Systematic Review and Meta-Analysis. J Am Heart Assoc 2022; 11(14): e025653.

14. Zuin M, Roncon L, Passaro A, Bosi C, Cervellati C, Zuliani G. Risk of dementia in patients with atrial fibrillation: Short versus long follow-up. A systematic review and meta-analysis. Int J Geriatr Psychiatry 2021; 36(10): 1488–500.

15. Papanastasiou CA, Theochari CA, Zareifopoulos N, et al. Atrial Fibrillation Is Associated with Cognitive Impairment, All-Cause Dementia, Vascular Dementia, and Alzheimer’s Disease: a Systematic Review and Meta-Analysis. J Gen Intern Med 2021; 36(10): 3122–35.

16. Proietti R, AlTurki A, Vio R, et al. The association between atrial fibrillation and Alzheimer’s disease: fact or fallacy? A systematic review and meta-analysis. J Cardiovasc Med (Hagerstown) 2020; 21(2): 106–12.

17. Saglietto A, Matta M, Gaita F, Jacobs V, Bunch TJ, Anselmino M. Stroke-independent contribution of atrial fibrillation to dementia: a meta-analysis. Open Heart 2019; 6(1): e000984.

18. Liu DS, Chen J, Jian WM, Zhang GR, Liu ZR. The association of atrial fibrillation and dementia incidence: a meta-analysis of prospective cohort studies. J Geriatr Cardiol 2019; 16(3): 298–306.

19. Kim D, Yang PS, Yu HT, et al. Risk of dementia in stroke-free patients diagnosed with atrial fibrillation: data from a population-based cohort. Eur Heart J 2019; 40(28): 2313–23.

20. Chen LYC, 2018 #75}, Norby FL, Gottesman RF, et al. Association of Atrial Fibrillation With Cognitive Decline and Dementia Over 20 Years: The ARIC-NCS (Atherosclerosis Risk in Communities Neurocognitive Study). J Am Heart Assoc 2018; 7(6).

21. de Bruijn RFAG, Heeringa J, Wolters FJ, et al. Association Between Atrial Fibrillation and Dementia in the General Population. JAMA Neurology 2015; 72(11): 1288–94.

22. Kalantarian S, Stern TA, Mansour M, Ruskin JN. Cognitive impairment associated with atrial fibrillation: a meta-analysis. Ann Intern Med 2013; 158(5 Pt 1): 338-46.

23. Islam MM, Poly TN, Walther BA, et al. Association Between Atrial Fibrillation and Dementia: A Meta-Analysis. Front Aging Neurosci 2019; 11: 305.

24. Santangeli P, Di Biase L, Bai R, et al. Atrial fibrillation and the risk of incident dementia: a meta-analysis. Heart Rhythm 2012; 9(11): 1761–8.

25. Alonso A, Knopman DS, Gottesman RF, et al. Correlates of Dementia and Mild Cognitive Impairment in Patients With Atrial Fibrillation: The Atherosclerosis Risk in Communities Neurocognitive Study (ARIC-NCS). J Am Heart Assoc 2017; 6(7).

26. Knecht S, Oelschläger C, Duning T, et al. Atrial fibrillation in stroke-free patients is associated with memory impairment and hippocampal atrophy. Eur Heart J 2008; 29(17): 2125–32.

27. Kühne M, Krisai P, Coslovsky M, et al. Silent brain infarcts impact on cognitive function in atrial fibrillation. Eur Heart J 2022; 43(22): 2127–35.

28. Silva DS, Caseli BG, de Campos BM, et al. Cerebral Structure and Function in Stroke-free Patients with Atrial Fibrillation. J Stroke Cerebrovasc Dis 2021; 30(8): 105887.

29. Berman JP, Norby FL, Mosley T, et al. Atrial Fibrillation and Brain Magnetic Resonance Imaging Abnormalities. Stroke 2019; 50(4): 783–8.

30. Piers RJ, Nishtala A, Preis SR, et al. Association between atrial fibrillation and volumetric magnetic resonance imaging brain measures: Framingham Offspring Study. Heart Rhythm 2016; 13(10): 2020–4.

31. Graff-Radford J, Madhavan M, Vemuri P, et al. Atrial fibrillation, cognitive impairment, and neuroimaging. Alzheimers Dement 2016; 12(4): 391–8.

32. Stefansdottir H, Arnar DO, Aspelund T, et al. Atrial fibrillation is associated with reduced brain volume and cognitive function independent of cerebral infarcts. Stroke 2013; 44(4): 1020–5.

33. Abe TA, Tressel W, Bartz TM, et al. Subclinical cardiac dysfunction and circulating markers of brain injury in older adults: The cardiovascular health study. J Stroke Cerebrovasc Dis 2025; 34(12): 108465.

34. Sudlow C, Gallacher J, Allen N, et al. UK biobank: an open access resource for identifying the causes of a wide range of complex diseases of middle and old age. PLoS Med 2015; 12(3): e1001779.

35. Alfaro-Almagro F, Jenkinson M, Bangerter NK, et al. Image processing and Quality Control for the first 10,000 brain imaging datasets from UK Biobank. Neuroimage 2018; 166: 400–24.

36. Miller KL, Alfaro-Almagro F, Bangerter NK, et al. Multimodal population brain imaging in the UK Biobank prospective epidemiological study. Nature Neuroscience 2016; 19(11): 1523–36.

37. Zhang Y, Brady M, Smith S. Segmentation of brain MR images through a hidden Markov random field model and the expectation-maximization algorithm. IEEE Trans Med Imaging 2001; 20(1): 45–57.

38. Patenaude B, Smith SM, Kennedy DN, Jenkinson M. A Bayesian model of shape and appearance for subcortical brain segmentation. Neuroimage 2011; 56(3): 907–22.

39. Griffanti L, Zamboni G, Khan A, et al. BIANCA (Brain Intensity AbNormality Classification Algorithm): A new tool for automated segmentation of white matter hyperintensities. Neuroimage 2016; 141: 191–205.

40. Cox SR, Ritchie SJ, Tucker-Drob EM, et al. Ageing and brain white matter structure in 3,513 UK Biobank participants. Nat Commun 2016; 7: 13629.

41. Fawns-Ritchie C, Deary IJ. Reliability and validity of the UK Biobank cognitive tests. PLoS One 2020; 15(4): e0231627.

42. Guo Y, You J, Zhang Y, et al. Plasma proteomic profiles predict future dementia in healthy adults. Nature Aging 2024; 4(2): 247–60.

43. Verberk IMW, Laarhuis MB, van den Bosch KA, et al. Serum markers glial fibrillary acidic protein and neurofilament light for prognosis and monitoring in cognitively normal older people: a prospective memory clinic-based cohort study. The Lancet Healthy Longevity 2021; 2(2): e87–e95.

44. Baskaran G, Krisai P, Kühne M, et al. Serum Neurofilament Light Chain and Cardiovascular Outcomes in Patients With Atrial Fibrillation. JAMA Cardiology 2026.

45. von Elm E, Altman DG, Egger M, Pocock SJ, Gøtzsche PC, Vandenbroucke JP. The Strengthening the Reporting of Observational Studies in Epidemiology (STROBE) statement: guidelines for reporting observational studies. BMJ 2007; 335(7624): 806-8.

46. Roselli C, Surakka I, Olesen MS, et al. Meta-analysis of genome-wide associations and polygenic risk prediction for atrial fibrillation in more than 180,000 cases. Nat Genet 2025; 57(3): 539–47.

47. Mishra A, Malik R, Hachiya T, et al. Stroke genetics informs drug discovery and risk prediction across ancestries. Nature 2022; 611(7934): 115–23.

48. Gallingani C, Carbone C, Tondelli M, Zamboni G. Neurofilaments Light Chain in Neurodegenerative Dementias: A Review of Imaging Correlates. Brain Sci 2024; 14(3).

49. Sjölin K, Aulin J, Wallentin L, et al. Serum Neurofilament Light Chain in Patients With Atrial Fibrillation. J Am Heart Assoc 2022; 11(14): e025910.

50. Silva DS, Coan AC, Avelar WM. Neuropsychological and neuroimaging evidences of cerebral dysfunction in stroke-free patients with atrial fibrillation: A review. J Neurol Sci 2019; 399: 172–81.

51. Moazzami K, Shao IY, Chen LY, et al. Atrial Fibrillation, Brain Volumes, and Subclinical Cerebrovascular Disease (from the Atherosclerosis Risk in Communities Neurocognitive Study [ARIC-NCS]). Am J Cardiol 2020; 125(2): 222–8.

52. Diener HC, Hart RG, Koudstaal PJ, Lane DA, Lip GYH. Atrial Fibrillation and Cognitive Function: JACC Review Topic of the Week. J Am Coll Cardiol 2019; 73(5): 612–9.

53. Manolis TA, Manolis AA, Apostolopoulos EJ, Melita H, Manolis AS. Atrial Fibrillation and Cognitive Impairment: An Associated Burden or Burden by Association? Angiology 2020; 71(6): 498–519.

54. Passey S, Patel J, Patail H, Aronow W. Association of Atrial Fibrillation and Cognitive Dysfunction: A Comprehensive Narrative Review of Current Understanding and Recent Updates. J Clin Med 2024; 13(18).

55. Zhai Y, Hu F, Yuan L, et al. Atrial fibrillation increases the risk of all-cause dementia, Alzheimer’s disease, and vascular dementia: A cohort study of 373, 415 participants in the UK Biobank. J Affect Disord 2024; 351: 323–30.

56. Ya-Nan O, Kevin K, Liu Y, et al. Longitudinal associations of cardiovascular health and vascular events with incident dementia. Stroke and Vascular Neurology 2024; 9(4): null.

57. Li M, Jiang C, Lai Y, et al. Genetic Evidence for Causal Association Between Atrial Fibrillation and Dementia: A Mendelian Randomization Study. Journal of the American Heart Association 2023; 12(16): e029623.

58. Junejo RT, Lip GYH, Fisher JP. Cerebrovascular Dysfunction in Atrial Fibrillation. Front Physiol 2020; 11: 1066.

59. Zenger B, Rizzi S, Steinberg BA, Ranjan R, Bunch TJ. This is Your Brain, and This is Your Brain on Atrial Fibrillation: The Roles of Cardiac Malperfusion Events and Vascular Dysfunction in Cognitive Impairment. Arrhythm Electrophysiol Rev 2023; 12: e01.

60. Chen LY, Agarwal SK, Norby FL, et al. Persistent but not Paroxysmal Atrial Fibrillation Is Independently Associated With Lower Cognitive Function: ARIC Study. J Am Coll Cardiol 2016; 67(11): 1379–80.

