## Supplementary Materials for "Atrial Fibrillation as a Determinant of Brain Health: Multimodal Evidence Supports Stroke-Dependent and Stroke-Independent Effects"

**eMethods**

*Imaging analysis*

*Cognitive function analyses*

*Dementia analyses*

*Protein Markers*

*Mendelian Randomisation*

*Genetic Instruments*

**Supplementary References**

**eTables**

**eTable 1:** ICD10, OPCS4 and verbal interview codes used to identify atrial fibrillation in the hospital diagnoses and procedures data (HES / SMR / PEDW) and the UK biobank verbal interview data.

**eTable 2:** ICD10 and verbal interview codes used to identify dementia in the hospital diagnoses and procedures data and the UK biobank verbal interview

**eTable 3:** Details of regression and adjustments for the analyses

**eTable 4:** ICD10 codes used to identify stroke in the hospital diagnoses and procedures data (HES/SMR/PEDW).

**eTable 5:** Phenotype definitions used to identify other cardiometabolic diseases in the hospital diagnoses and procedures data and the UK biobank verbal interview data.

**eTable 6:** Association between genetically proxied atrial fibrillation and cognitive tests at the second imaging visit

**eTable 7:** Characteristics of the dementia cohort (individuals aged over 65 at any point during follow-up, with no record of dementia before their 65th birthday)

**eTable 8:** The 269 SNPs retained as the AF instrument, the associated summary statistics and the summary statistics for ischaemic stroke (GIGASTROKE)

**eFigures**

**eFigure 1:** Study design showing the number of individuals in each analysis and the associated exclusions.

**eFigure 2:** The association between AF (n=2538) and regional brain volume IDPs in 72 073 individuals (a) adjusted for demographic factors and (b) fully adjusted.

**eFigure 3:** Fully adjusted association of (a) recruitment NFL or (b) recruitment GFAP with prevalent AF and ischaemic stroke

#### eMethods

*Imaging analysis*

The neuroimaging data acquisition and pre-processing pipelines have been described previously [1 2]. In brief, the data used in this project were acquired at three imaging centres using identical 3 T Siemens Skyra MRI scanners (software VD13) and the Siemens 32-Channel Head Coil. The scanning protocol consisted of structural T1-weighted MRI (T1), resting-state functional MRI, task fMRI, T2-weighted fluid-attenuated inversion recovery (FLAIR) imaging, diffusion MRI (dMRI) and susceptibility-weighted MRI. Measurements were taken from 2017 onwards. For this paper, we analysed data from the T1, T2, and dMRI modalities, which provide information on GM structure (T1), volumes of white matter hyperintensities (FLAIR), and WM structure/connectivity (dMRI).

The UK Biobank (UKB) has developed automated pre-processing pipelines for imaging data that provide a large number of image-derived phenotypes (IDPs), which were used in this paper as provided. For the T1-weighted data, the IDPs analysed in this study included FAST-derived subcortical volumes (caudate, pallidum, amygdala, putamen, hippocampus, thalamus, accumbens) and FIRST-derived GM volumes not including the grey matter in the subcortical regions (caudate, pallidum, amygdala, putamen, hippocampus, thalamus). Some of these subcortical areas may be affected by the issue of white matter hyperintensities being misidentified as grey matter in the automated pipeline, leading to erroneous positive associations between stroke and the regional grey matter volume [3 4]. All volumetric IDPs were normalised for head size prior to analysis, and WMH volume was log-transformed.

The IDPs used from diffusion MRI were fractional anisotropy (FA), mean diffusivity (MD). Pre-processed diffusion MRI IDPs included the weighted means of the FA and MD across 27 white matter tracts. These values are strongly correlated, and for simplicity, we created a single metric of FA and MD, the global FA and global MD created through factor analysis [5].

Individuals were included in the imaging analysis if they had complete FIRST/FAST and diffusion IDPs, and complete imaging covariates. Individuals with excessive motion or an excessive number of diffusion outliers, defined as resting-function mean head motion > 3 standard deviations from the mean resting-function mean head motion or the number of diffusion outliers > 3 standard deviations from the mean number of diffusion outliers respectively, were excluded. A total of 54 601 individuals were analysed.

Linear regression was used to analyse the association between AF and the brain imaging IDPs. All analyses were adjusted for age, $age^2$, sex, age*sex, $age^2$*sex, and imaging-derived confounders[6]. Imaging confounders included the volumetric scaling factor (for non-volumetric IDPs), head motion (linear and quadratic) from the resting state function MRI, head position (x,y,z) in the MRI scanner (linear and quadratic), month of scan (categorical) to account for temporal drifts in the data, imaging site and interaction between imaging site and the other imaging confounders. Other covariates are listed in **eTable 3a**. Individuals were treated as having a coexisting disease (diabetes, heart failure, coronary heart disease, non-haemorrhagic stroke or ischaemic stroke) if the disease was first reported on or before the date of the imaging visit.

For the analysis of the regional volume IDPs, the false discovery rate due to multiple testing was controlled for by the method of Benjamini-Yekutieli (BY). Results were considered significant if BY-adjusted p-values were <0.05. For the seven global IDPs, significance was assessed using a Bonferroni-corrected threshold of $\alpha = 0.05/7 = 0.007$. The global IDPs are hightly correlated, so this is likely to be conservative.

*Cognitive function analyses*

For each cognitive test, linear regression was used to analyse the association between pre-assessment AF or stroke and the cognitive function score (in units of 1 standard deviation). Analyses were adjusted for age at the time of the test (single year categorical variable), and other covariates listed in **eTable 3a**. Individuals were considered to have coexisting diseases if they were first recorded on or before the date of the cognitive assessment.

For analysis of the association of AF with the five cognitive test results, significance was assessed using a Bonferroni-corrected threshold of $\alpha = 0.05/5 = 0.01$.

*Dementia analyses*

Cox regression, with age-at-risk as the time scale and stratified by recruitment year, was used to investigate the association between a first report of AF or stroke before age 65 and a first report of dementia after age 65. The chance of finding a dementia record in the HADP data is greater for individuals with multiple hospital admissions. To account for this, the analyses were also stratified by grouped hospitalisations per year over age 65 (counting only hospitalisations where dementia was not the primary diagnosis). Prevalent and incident coexisting diseases were included as time-dependent covariates (0 before first report, 1 afterwards). More details about covariates are given in **eTable 3b**.

For analysis of the association of AF with the dementia endpoints, significance was assessed using a Bonferroni-corrected threshold of $\alpha = 0.05/4 = 0.0125$.

*Protein Markers*

Plasma protein expression for a large number of proteins, including plasma neurofilament light chain (NfL) and glial fibrillary acidic protein (GFAP), was measured in the UK biobank in around 50 000 individuals at recruitment using the Olink target platform [7].

After excluding individuals who were not part of the random selection, 44 369 individuals had measured values for NfL and 44 339 individuals had measured values for GFAP. These included 732 individuals with pre-recruitment AF (self-reported or hospital episode-sourced) and 242 individuals with pre-recruitment stroke (hospital episode-sourced). Olink protein concentrations are reported on a $\log_2$ scale in NPX units. Thus, a difference in 1 NPX represents a doubling (100% increase) in protein concentration. To convert a change of δ in NPX units to a percent change, we used percent change=$100\times(2^{\delta}-1)$.

Linear regression was used to study the mutually adjusted association between pre-recruitment AF or stroke and plasma NfL and GFAP expression. The models included both pre-recruitment AF and ischaemic stroke and analyses were further adjusted for baseline age (linear and quadratic), sex, sex*age, plate, Townsend deprivation index and education, recruitment visit smoking status, systolic blood pressure, body mass index, grouped alcohol consumption, HDL-, LDL-, and total cholesterol, prevalent heart failure, coronary heart disease and diabetes (**eTable 3b)**.

*Mendelian randomisation*

To investigate whether any observed AF–outcome associations could be mediated by ischaemic stroke, we conducted univariate two-sample Mendelian randomisation (MR) using an AF genetic instrument to estimate the total effect of genetically-proxied AF on each outcome, plus multivariate MR to examine the associations conditional on ischaemic stroke.

*Genetic instruments*

The genetic instrument for AF was based on variants reported in the recent GWAS meta-analysis of Roselli *et al.*[8], which included more than 180,000 AF cases. We restricted to SNPs with a p-value $< 5 \times 10^{-8}$ that overlapped with the UK Biobank genotyping panel and had available association estimates in the GIGASTROKE consortium summary statistics[9]. Variants were then clumped using the *ieugwasr* R package[10]. Clumping was performed with a physical distance threshold of 10,000 kilobases, such that only the most significant SNP was retained within each 10 Mb window, and an LD threshold of $r^2$=0.001, and a liberal p-value threshold of 0.99. Effect alleles were harmonised across datasets to ensure alignment. The SNP–AF associations used in this instrument represent log-odds of AF per effect allele estimated from logistic regression in the discovery GWAS. After quality control and harmonisation, this process yielded 269 independent SNPs that formed the AF genetic instrument (**eTable 7**).

Summary statistics for the outcomes in our primary analysis were obtained by regressing the same 269 SNPs on the outcome in the UKB dataset. For the multivariate MR, summary statistics for ischaemic stroke for the same 269 SNPs were sourced from the *GIGASTROKE* consortium[9].

All analyses were performed using SAS v9.4 and R v4.4.2.

**Supplementary References**


1. Miller KL, Alfaro-Almagro F, Bangerter NK, et al. Multimodal population brain imaging in the UK Biobank prospective epidemiological study. *Nature Neuroscience* 2016; **19**(11): 1523-36.
2. Alfaro-Almagro F, Jenkinson M, Bangerter NK, et al. Image processing and Quality Control for the first 10,000 brain imaging datasets from UK Biobank. *Neuroimage* 2018; **166**: 400-24.
3. Dadar M, Potvin O, Camicioli R, Duchesne S. Beware of white matter hyperintensities causing systematic errors in FreeSurfer gray matter segmentations! *Hum Brain Mapp* 2021; **42**(9): 2734-45.
4. Griffanti L, Gillis G, O'Donoghue MC, et al. Adapting UK Biobank imaging for use in a routine memory clinic setting: The Oxford Brain Health Clinic. *NeuroImage: Clinical* 2022; **36**: 103273.
5. Cox SR, Ritchie SJ, Tucker-Drob EM, et al. Ageing and brain white matter structure in 3,513 UK Biobank participants. *Nat Commun* 2016; **7**: 13629.
6. Alfaro-Almagro F, McCarthy P, Afyouni S, et al. Confound modelling in UK Biobank brain imaging. *NeuroImage* 2021; **224**: 117002.
7. Sun BB, Chiou J, Traylor M, et al. Genetic regulation of the human plasma proteome in 54,306 UK Biobank participants. *bioRxiv* 2022: 2022.06.17.496443.
8. Roselli C, Surakka I, Olesen MS, et al. Meta-analysis of genome-wide associations and polygenic risk prediction for atrial fibrillation in more than 180,000 cases. *Nat Genet* 2025; **57**(3): 539-47.
9. Mishra A, Malik R, Hachiya T, et al. Stroke genetics informs drug discovery and risk prediction across ancestries. *Nature* 2022; **611**(7934): 115-23.

10. Hemani G, Elsworth B, Palmer T, Rasteiro R (2025). ieugwasr: Interface to the 'OpenGWAS' Database API. R package version 1.1.0, https://github.com/MRCIEU/ieugwasr.

**eTable 1: ICD10, OPCS4 and verbal interview codes used to identify atrial fibrillation in the hospital diagnoses and procedures data (HES / SMR / PEDW) and the UK biobank verbal interview data.**

| Code Value | Condition |
|---|---|
| **ICD10 Codes** | |
| I48 | Atrial fibrillation and flutter |
| I480 | Paroxysmal atrial fibrillation |
| I481 | Persistent atrial fibrillation |
| I482 | Chronic atrial fibrillation |
| I483 | Typical atrial flutter |
| I484 | Atypical atrial flutter |
| I489 | Atrial fibrillation and atrial flutter, unspecified |
| **OPCS4 Codes** | |
| K223 | Exclusion of left atrial appendage NEC |
| K571 | Percutaneous transluminal ablation of atrioventricular node |
| K575 | Percutaneous transluminal ablation of atrial wall NEC |
| K621 | Percutaneous transluminal ablation of pulmonary vein to left atrium conducting system |
| K622 | Percutaneous transluminal ablation of atrial wall for atrial flutter |
| K623 | Percutaneous transluminal ablation of conducting system of heart for atrial flutter NEC |
| K624 | Percutaneous transluminal internal cardioversion NEC |
| K625 | Percutaneous transluminal occlusion of left atrial appendage |
| X501 | Direct current cardioversion |
| X502 | External cardioversion NEC |
| **UKB Verbal interview codes (self-reported)** | |
| **Non-cancer illness code, UKB field id=20002** | |
| 1471 | Atrial Fibrillation |
| 1483 | Atrial Flutter |
| **Operation codes, UKB field id=20004** | |
| 1553 | Cardiac Ablation |
| 1524 | Cardioversion |

**eTable 2: ICD10 and verbal interview codes used to identify dementia in the hospital diagnoses and procedures data and the UK biobank verbal interview**

| Code Value | Condition |
|---|---|
| **ICD10 Codes** | |
| | **Alzheimer's disease:** |
| F00 | Dementia in Alzheimer's disease |
| F00.0 | Dementia in Alzheimer's disease with early onset |
| F00.1 | Dementia in Alzheimer's disease with late onset |
| F00.2 | Dementia in Alzheimer's disease, atypical or mixed type |
| F00.9 | Dementia in Alzheimer's disease, unspecified |
| G30 | Alzheimer’s disease |
| G30.0 | Alzheimer’s disease with early onset |
| G30.1 | Alzheimer’s disease with late onset |
| G30.8 | Other Alzheimer's disease |
| G30.9 | Alzheimer's disease unspecified |
| | **Vascular dementia:** |
| F01 | Vascular dementia |
| F01.0 | Vascular dementia of acute onset |
| F01.1 | Multi-infarct dementia |
| F01.2 | Subcortical vascular dementia |
| F01.3 | Mixed cortical and sub-cortical vascular dementia |
| F01.8 | Other vascular dementia |
| F01.9 | Vascular dementia, unspecified |
| I67.3 | Binswanger's disease |
| | **Frontotemporal dementia:** |
| F02.0 | Dementia in Picks disease |
| G31.0 | Circumscribed brain atrophy |
| | **Other and unclassified dementia:** |
| A81.0 | Sporadic Creutzfeldt-Jakob disease |
| F02 | Dementia in other diseases classified elsewhere |
| F02.1 | Dementia in Creutzfeldt-Jacob disease |
| F02.2 | Dementia in Huntington’s disease |
| F02.3 | Dementia in Parkinson’s disease |
| F02.4 | Dementia in HIV disease |
| F02.8 | Dementia in other specified diseases classified elsewhere |
| F03 | Unspecified dementia |
| F05.1 | Delirium superimposed on dementia |
| F10.6 | Mental and behavioural disorders due to use of alcohol - amnesic syndrome |
| G31.1 | Senile degeneration of brain |
| G31.8 | Other specified degenerative diseases of nervous system |
| **UKB Verbal interview codes (self-reported)** | |
| **Non-cancer illness code, UKB field id=20002** | |
| 1263 | Dementia/Alzheimers/Cognitive Impairment |

**eTable 3a: Details of regression and adjustments for the analysis of the association between diagnosed AF and cognitive outcomes - brain imaging markers and cognitive scores**

| Outcome | Exposure | Regression | Covariates for minimally adjusted analysis | Additional covariates for full analysis |
|---|---|---|---|---|
| Brain IDPs (SD units) | AF diagnosis first recorded before imaging visit | Linear | age_at_imaging, age_at_imaging$^2$<br>sex, age_at_imaging*sex, age_at_imaging$^2$*sex<br>Highest education level[f]<br>Recruitment Townsend deprivation index[d]<br>Imaging covariates:<br>volumetric scaling factor [for non-volumetric IDPs], head motion (linear and quadratic) from the resting state function MRI, head position (x,y,z) in the MRI scanner (linear and quadratic), month of scan (categorical) to account for temporal drifts in the data, imaging site and interaction between imaging site and the other imaging confounders. | *CVD risk factors:*<br>Recruitment visit smoking status[a]<br>Grouped alcohol consumption[b]<br>Grouped body mass index[c]<br>Grouped systolic blood pressure[c]<br>LDL-, HDL- and total cholesterol[d]<br>diabetes recorded prior to imaging visit (0/1)<br>*Other cardiovascular comorbidities[e]:*<br>Heart failure prior to imaging visit (0/1)<br>Coronary heart disease prior to imaging visit (0/1) |
| Cognitive score (SD units) | AF diagnosis first recorded before cognitive assessment | Linear | age at test (single year, categorical)<br>sex<br>Highest education level[f]<br>Recruitment Townsend deprivation index[d] | *CVD risk factors:*<br>Recruitment visit smoking status[a]<br>Grouped alcohol consumption[b]<br>Grouped body mass index[c]<br>Grouped systolic blood pressure[c]<br>LDL-, HDL- and total cholesterol[d]<br>diabetes recorded prior to cognitive assessment (0/1)<br>*Other cardiovascular comorbidities[e]:*<br>Heart failure prior to cognitive assessment (0/1)<br>Coronary heart disease prior to cognitive assessment (0/1)<br>Haemorrhagic stroke prior to cognitive assessment (0/1)<br>Ischaemic stroke prior to cognitive assessment (0/1) |

[a] Categorical variable (never: current :ex-: unknown). [b] Categorical variable grouped by units/week (<3: 3 - <8 : 8 - <16: >16 : unknown) [c] Quintilesor unknown. [d] Continuous with binary missing indicator [e] Definitions of other cardiomorbidities are given in eTables 4 and 7. [f] Categorical variable (degree level: A-level or equivalent: none of these/missing)

### eTable 3b: Details of regression and adjustments for the analysis of the association between diagnosed AF and cognitive outcomes - incident dementia and recruitment proteins

| Outcome | Exposure | Regression | Covariates for minimally adjusted analysis | Additional covariates for full analysis |
|---|---|---|---|---|
| Dementia first recorded after age 65 | AF diagnosis before age 65 | Cox regression with age at risk as the timescale | Stratified by mean hospitalisations/year over age 65 (not including admissions where dementia is the primary diagnosis.<br><br>Stratified by individual year of recruitment<br>*Covariates:*<br>Sex<br>Highest education level[f]<br>Recruitment Townsend deprivation index[d] | *CVD risk factors:*<br>Recruitment visit smoking status[a]<br>Grouped alcohol consumption[b]<br>Grouped body mass index[c]<br>Grouped systolic blood pressure[c]<br>LDL-, HDL- and total cholesterol[d]<br>prevalent and incident diabetes (time-dependent covariate)<br>*Other cardiovascular comorbidities[e] (time-dependent covariates, 0 before first report, 1 afterwards):*<br>prevalent and incident heart failure<br>prevalent and incident coronary heart disease<br>prevalent and incident haemorrhagic stroke<br>prevalent and incident ischaemic stroke |
| neurofilament light chain or glial fibrillary acidic protein | Pre-recruitment record of AF and, in a joint analysis, pre-recruitment record of ischaemic stroke | Linear | n/a | age_at_recruitment, age_at_recruitment$^2$<br>sex, age_at_recruitment*sex, plate<br>Highest education level[e]<br>Recruitment Townsend deprivation index[c]<br>Recruitment visit smoking status[a]<br>Grouped alcohol consumption[b]<br>Body mass index[c]<br>Systolic blood pressure[c]<br>LDL-, HDL- and total cholesterol[d]<br>Pre-recruitment heart failure, coronary heart disease and diabetes[e] (0/1)<br>[eGFR added as a sensitivity analysis] |

[a] Categorical variable (never: current :ex-: unknown). [b] Categorical variable grouped by units/week (<3: 3 - <8 : 8 - <16: >16 : unknown) [c] Quintilesor unknown. [d] Continuous with binary missing indicator [e] Definitions of other cardiomorbidities are given in eTables 4 and 7. [f] Categorical variable (degree level: A-level or equivalent: none of these/missing)

### eTable 4: ICD10 codes used to identify stroke in the hospital diagnoses and procedures data (HES/SMR/PEDW).

| Code Value | Condition |
|---|---|
| *Stroke not known to be haemorragic* | |
| I63 | Cerebral infarction |
| I630 | Cerebral infarction due to thrombosis of precerebral arteries |
| I631 | Cerebral infarction due to embolism of precerebral arteries |
| I632 | Cerebral infarction due to unspecified occlusion or stenosis of precerebral arteries |
| I633 | Cerebral infarction due to thrombosis of cerebral arteries |
| I634 | Cerebral infarction due to embolism of cerebral arteries |
| I635 | Cerebral infarction due to unspecified occlusion or stenosis of cerebral arteries |
| I636 | Cerebral infarction due to cerebral venous thrombosis, nonpyogenic |
| I638 | Other cerebral infarction |
| I639 | Cerebral infarction, unspecified |
| I64 | Stroke, not specified as haemorrhage or infarction |
| *Haemorrhagic stroke* | |
| I60 | Subarachnoid haemorrhage |
| I600 | Subarachnoid haemorrhage from carotid siphon and bifurcation |
| I601 | Subarachnoid haemorrhage from middle cerebral artery |
| I602 | Subarachnoid haemorrhage from anterior communicating artery |
| I603 | Subarachnoid haemorrhage from posterior communicating artery |
| I604 | Subarachnoid haemorrhage from basilar artery |
| I605 | Subarachnoid haemorrhage from vertebral artery |
| I606 | Subarachnoid haemorrhage from other intracranial arteries |
| I607 | Subarachnoid haemorrhage from intracranial artery, unspecified |
| I608 | Other subarachnoid haemorrhage |
| I609 | Subarachnoid haemorrhage, unspecified |
| I61 | Intracerebral haemorrhage |
| I610 | Intracerebral haemorrhage in hemisphere, subcortical |
| I611 | Intracerebral haemorrhage in hemisphere, cortical |
| I612 | Intracerebral haemorrhage in hemisphere, unspecified |
| I613 | Intracerebral haemorrhage in brain stem |
| I614 | Intracerebral haemorrhage in cerebellum |
| I615 | Intracerebral haemorrhage, intraventricular |
| I616 | Intracerebral haemorrhage, multiple localised |
| I618 | Other intracerebral haemorrhage |
| I619 | Intracerebral haemorrhage, unspecified |
| I62 | Other nontraumatic intracranial haemorrhage |
| I620 | Subdural haemorrhage (acute) (nontraumatic) |
| I621 | Nontraumatic extradural haemorrhage |
| I629 | Intracranial haemorrhage (nontraumatic), unspecified |

Note that ischaemic stroke includes stroke not known to be haemorrhagic

**eTable 5: Phenotype definitions used to identify other cardiometabolic diseases in the hospital diagnoses and procedures data and the UK biobank verbal interview data.**

| | |
|---|---|
| **Heart failure** | |
| ICD10 codes | I50 ,I50.0, I50.1, I50.9,<br>I11.0, I13.0 ,I13.2,<br>I25.5,<br>I42.0, I42.3, I42.4, I42.5, I42.6, I42.7, I42.8, I42.9,<br>I43, I43.0, I43.1, I43.2, I43.8, |
| OPCS4 codes | K54.1, K54.8, K54.9,<br>K56.2, K56.3, K56.8, K56.9,<br>K60.7. K61.7, K73.3, K74.3,<br>K01.1, K01.2, K01.8, K01.9,<br>K02.1, K02.2, K02.3, K02.4, K02.5, K02.6, K02.8, K02.9 |
| UKB self report | |
| Non-Cancer illnesses (field 20002) | 1076 (heart failure or pulmonary odema);<br>1079 (Cardiomyopathy) |
| **Coronary heart disease** | |
| ICD10 codes | I20, I20.0, I20.1, I20.8, I20.9,<br>I21, I21.0, I21.1, I21.2, I21.3, I21.4, I21.9,<br>I22, I22.0, I22.1, I22.8, I22.9,<br>I23, I23.0, I23.1, I23.2, I23.3, I23.4, I23.5, I23.6, I23.8,<br>I24, I24.0, I24.1, I24.8, I24.9,<br>I25, I25.0, I25.1, I25.2, I25.3, I25.4, I25.6, I25.8, I25.9 |
| OPCS4 codes | K49, K49.1, K49.2, K49.3, K49.4, K49.8, K49.9,<br>K50, K50.1, K50.2, K50.3, K50.4, K50.8, K50.9,<br>K75, K75.1, K75.2, K75.3, K75.4, K75.8, K75.9,<br>K40., K40.1, K40.2, K40.3, K40.4, K40.8, K40.9,<br>K41., K41.1, K41.2, K41.3, K41.4, K41.8, K41.9,<br>K42., K42.1, K42.2, K42.3, K42.4, K42.8, K42.9,<br>K43., K43.1, K43.2, K43.3, K43.4, K43.8, K43.9,<br>K44., K44.1, K44.2, K44.8, K44.9,<br>K45., K45.1, K45.2, K45.3, K45.4, K45.5, K45.6, K45.8, K45.9,<br>K46., K46.1, K46.2, K46.3, K46.4, K46.5, K46.8, K46.9, |
| UKB self report | |
| Non-Cancer illnesses (field 20002) | 1075 (Heart attack/myocardial infarction) |
| Operations (field 20004) | 1070 (Coronary angioplasty (PTCA) +/- stent);<br>1095 (Coronary artery bypass grafts)<br>1523 (Triple heart bypass) |
| **Diabetes** | |
| ICD10 codes | E10, E100, E101, E102, E103, E104, E105, E106, E107, E108, E109, E11, E110, E112, E113, E114, E115, E116, E118, E119, E14, E140, E142, E143, E144, E145, E146, E147, E148, E149, E13, E130, E131, E132, E133, E134, E135, E136, E137, E138, E139 |
| UKB self-report | |
| Non-Cancer illnesses (field 20002) | 1220, 1222, 1223 |
| Diabetes specific medication | 1140883066<br>1140868902, 1141171652, 1141156984, 1141189094, 1140874724, 1140874736, 1140874746, 1140874718, 1140874650, 1140874744, 1141152590, 1140874646, 1141157284, 1140874646, 1140874686, 1140910566, 1140883066, 1140884600, 1140874652, 1141173882, 1141168668, 1141171646, 1141168660, 1141177600, 1141189090, 1141173786, 1140874726, 1140874674, 1140874666 |
| Diabetes diagnosed by a doctor | field 2443 |
| Glycated hemoglobin (field-id 30750) | >=48 and <184 mmol/mol |

**eTable 6: Results of univariate inverse variance weighted MR for the association of genetically proxied AF with cognitive function test results from the UKB second online cognitive assessment**

| Cognitive Test | N[1] | Estimate (SE)[2] | P-value |
|---|---|---|---|
| Fluid intelligence | 131,349 | -0.008 (0.005) | 0.15 |
| Digit-symbol substitution | 128,972 | 0.000 (0.006) | 0.96 |
| Matrix pattern completion | 127,671 | -0.002 (0.006) | 0.73 |
| Trail making (trail 1) | 127,671 | 0.002 (0.006) | 0.74 |
| Trail making (trail 2) | 127,671 | -0.008 (0.006 | 0.17 |

[1] Number of European non-related individuals with genetic information and non-missing values
[2] units of standard deviation of score per unit increase in log odds of atrial fibrillation.

**eTable 7: Characteristics of the dementia cohort (individuals aged over 65 at any point during follow-up, with no record of dementia before their 65th birthday)**

| | All (N=365879) | With AF before 65 (N= 10462) |
|---|---|---|
| Age (years), mean (SD) | | |
| At recruitment | 60.5 (5.2) | 60.0 (5.2) |
| At start of post-65 follow-up | 65.6 (1.3) | 65.5 (1.2) |
| Female sex, n(%) | 198896 (54.4%) | 3423 (32.7%) |
| Mean follow-up, mean (SD) | 7.8 (4.3) | 6.2 (4.4) |
| Hospitalisation/year post-65 | | |
| Mean (SD) | 0.7 (2.2) | 1.3 (4.3) |
| 0, n(%) | 104663 (28.6%) | 1894 (18.1%) |
| <0.5, n(%) | 150472 (41.1%) | 3177 (30.4%) |
| 0.5-1, n(%) | 57322 (15.7%) | 2225 (21.3%) |
| >1, n(%) | 53422 (14.6%) | 3166 (30.3%) |
| Incident Dementia post-65, n(%) | | |
| Alzheimers | 3388 (0.9%) | 94 (0.9%) |
| Vascular dementia | 1507 (0.4%) | 83 (0.8%) |
| Frontotemporal dementia | 171 (0.0%) | 3 (0.0%) |
| Unspecified / other | 5115 (1.4%) | 213 (2.0%) |
| Any dementia | 10181 (2.8%) | 393 (3.8%) |

Abbreviations: SD=Standard deviation

**eTable 8: The 269 SNPs retained as the AF instrument, the associated summary statistics and the summary statistics for ischaemic stroke (GIGASTROKE**

| SNP ID | Chomosome: Position | UKB effect/ other allele | UKB effect allele freq | Atrial Fibrillation β (SE) | Atrial Fibrillation P-value | Ischaemic Stroke β (SE) | Ischaemic Stroke P-value |
|---|---|---|---|---|---|---|---|
| rs75350262 | 1:10424375 | G/A | 0.98 | -0.113 (0.016) | $1.1x10^{-12}$ | -0.010 (0.028) | 0.74 |
| rs880315 | 1:10736809 | T/C | 0.66 | -0.043 (0.005) | $7.4x10^{-18}$ | -0.045 (0.008) | $2.0x10^{-9}$ |
| rs6426798 | 1:19393234 | C/T | 0.72 | -0.029 (0.005) | $2.1x10^{-9}$ | -0.003 (0.008) | 0.67 |
| rs7529220 | 1:21956126 | T/C | 0.14 | -0.055 (0.006) | $1.1x10^{-20}$ | -0.013 (0.010) | 0.21 |
| rs61750827 | 1:38918953 | C/T | 0.94 | -0.066 (0.009) | $1.4x10^{-12}$ | -0.031 (0.015) | 0.039 |
| rs2885697 | 1:41078607 | G/T | 0.33 | 0.036 (0.005) | $8.1x10^{-13}$ | 0.000 (0.007) | 0.97 |
| rs72690501 | 1:50851367 | C/A | 0.98 | -0.193 (0.015) | $4.7x10^{-37}$ | -0.026 (0.023) | 0.27 |
| rs7543039 | 1:99584092 | C/T | 0.50 | -0.031 (0.004) | $4.1x10^{-12}$ | -0.019 (0.007) | $7.3x10^{-3}$ |
| rs10783115 | 1:99669780 | G/T | 0.22 | -0.031 (0.005) | $3.3x10^{-9}$ | 0.006 (0.009) | 0.51 |
| rs2813865 | 1:111895334 | A/G | 0.23 | -0.045 (0.005) | $2.1x10^{-17}$ | -0.009 (0.009) | 0.31 |
| rs4074536 | 1:115768346 | T/C | 0.70 | 0.058 (0.005) | $7.8x10^{-36}$ | -0.004 (0.010) | 0.69 |
| rs78581286 | 1:147844695 | G/A | 0.95 | 0.107 (0.010) | $2.1x10^{-29}$ | 0.029 (0.015) | 0.055 |
| rs6689306 | 1:154423470 | A/G | 0.42 | 0.045 (0.004) | $1.1x10^{-24}$ | 0.014 (0.007) | 0.057 |
| rs34515871 | 1:154839804 | C/T | 0.69 | -0.127 (0.005) | $6.5x10^{-154}$ | -0.006 (0.008) | 0.41 |
| rs4845408 | 1:155093151 | G/A | 0.84 | 0.046 (0.006) | $1.9x10^{-14}$ | 0.003 (0.009) | 0.73 |
| rs4987408 | 1:169689997 | T/C | 0.96 | -0.071 (0.012) | $4.9x10^{-9}$ | -0.021 (0.020) | 0.28 |
| rs72700118 | 1:170225682 | C/A | 0.88 | -0.123 (0.007) | $1.7x10^{-72}$ | -0.015 (0.011) | 0.17 |
| rs680084 | 1:170659114 | G/A | 0.46 | 0.088 (0.004) | $7.8x10^{-94}$ | 0.018 (0.007) | 0.012 |
| rs7520192 | 1:170681513 | A/G | 0.65 | -0.040 (0.005) | $5.1x10^{-19}$ | -0.002 (0.008) | 0.84 |
| rs11579055 | 1:203062187 | G/T | 0.45 | 0.073 (0.004) | $1.9x10^{-63}$ | 0.027 (0.007) | $2.0x10^{-4}$ |
| rs12116645 | 1:204555346 | G/T | 0.34 | -0.029 (0.005) | $3.3x10^{-9}$ | -0.025 (0.013) | 0.058 |
| rs2793374 | 1:205678380 | C/T | 0.53 | 0.033 (0.004) | $1.4x10^{-13}$ | -0.003 (0.007) | 0.69 |
| rs12023502 | 1:217183138 | C/T | 0.74 | -0.030 (0.005) | $2.4x10^{-10}$ | -0.002 (0.008) | 0.85 |
| rs145390113 | 1:227570293 | T/C | 0.97 | 0.084 (0.013) | $1.2x10^{-10}$ | -0.008 (0.020) | 0.71 |
| rs62107261 | 2:422144 | T/C | 0.95 | 0.064 (0.012) | $4.8x10^{-8}$ | -0.032 (0.020) | 0.10 |
| rs2867131 | 2:610603 | T/C | 0.17 | -0.037 (0.006) | $3.8x10^{-10}$ | -0.003 (0.010) | 0.74 |
| rs4396680 | 2:10038109 | A/G | 0.20 | 0.042 (0.006) | $5.2x10^{-14}$ | 0.010 (0.010) | 0.31 |
| rs11689727 | 2:25235231 | C/A | 0.68 | 0.027 (0.005) | $7.2x10^{-9}$ | -0.004 (0.008) | 0.63 |
| rs6546620 | 2:25937071 | T/C | 0.19 | -0.056 (0.006) | $1.3x10^{-23}$ | -0.002 (0.009) | 0.83 |

| SNP ID | Chomosome: Position | UKB effect/ other allele | UKB effect allele freq | Atrial Fibrillation β (SE) | Atrial Fibrillation P-value | Ischaemic Stroke β (SE) | Ischaemic Stroke P-value |
|---|---|---|---|---|---|---|---|
| rs12328620 | 2:37870755 | G/A | 0.60 | -0.029 (0.004) | $1.1x10^{-10}$ | -0.019 (0.008) | 0.013 |
| rs1867785 | 2:46307199 | A/G | 0.41 | 0.029 (0.004) | $1.5x10^{-10}$ | 0.004 (0.007) | 0.59 |
| rs4672423 | 2:61208140 | C/T | 0.62 | 0.038 (0.004) | $4.8x10^{-18}$ | 0.009 (0.007) | 0.21 |
| rs2723064 | 2:65052671 | T/C | 0.62 | 0.070 (0.004) | $3.4x10^{-56}$ | 0.016 (0.007) | 0.026 |
| rs10865380 | 2:69864114 | T/C | 0.54 | 0.048 (0.004) | $6.8x10^{-29}$ | 0.006 (0.007) | 0.42 |
| rs55945133 | 2:86344544 | C/T | 0.88 | 0.057 (0.008) | $1.0x10^{-13}$ | 0.018 (0.011) | 0.11 |
| rs3792130 | 2:99598153 | C/G | 0.17 | 0.035 (0.006) | $3.8x10^{-8}$ | 0.011 (0.010) | 0.28 |
| rs4663039 | 2:126665499 | G/A | 0.91 | -0.067 (0.008) | $2.2x10^{-19}$ | -0.037 (0.015) | 0.014 |
| rs6430286 | 2:148076795 | G/A | 0.58 | -0.030 (0.004) | $5.9x10^{-12}$ | -0.011 (0.007) | 0.15 |
| rs7574892 | 2:174648092 | G/A | 0.52 | -0.058 (0.004) | $5.5x10^{-42}$ | 0.013 (0.007) | 0.072 |
| rs890578 | 2:178549906 | C/A | 0.83 | -0.095 (0.006) | $6.8x10^{-65}$ | -0.029 (0.009) | $1.8x10^{-3}$ |
| rs6716898 | 2:198079547 | G/A | 0.51 | 0.031 (0.004) | $1.5x10^{-12}$ | 0.005 (0.007) | 0.44 |
| rs7605146 | 2:200319165 | G/A | 0.61 | -0.059 (0.004) | $1.2x10^{-41}$ | -0.002 (0.007) | 0.74 |
| rs13019524 | 2:212386779 | T/C | 0.67 | 0.041 (0.005) | $5.9x10^{-19}$ | -0.001 (0.008) | 0.91 |
| rs7589956 | 2:237624789 | C/T | 0.44 | -0.029 (0.004) | $4.1x10^{-11}$ | -0.011 (0.007) | 0.11 |
| rs6767174 | 3:12667015 | A/G | 0.68 | -0.026 (0.005) | $2.5x10^{-8}$ | 0.002 (0.007) | 0.75 |
| rs7650482 | 3:12800305 | A/G | 0.35 | -0.081 (0.005) | $7.4x10^{-72}$ | -0.002 (0.008) | 0.78 |
| rs779029 | 3:21915732 | A/C | 0.77 | 0.032 (0.006) | $1.3x10^{-8}$ | -0.000 (0.009) | 1.00 |
| rs73041705 | 3:24421744 | T/C | 0.70 | 0.048 (0.005) | $1.2x10^{-24}$ | 0.001 (0.008) | 0.95 |
| rs6808550 | 3:25066375 | A/G | 0.43 | -0.024 (0.004) | $3.3x10^{-8}$ | 0.006 (0.007) | 0.43 |
| rs6599219 | 3:38596219 | G/A | 0.34 | -0.036 (0.005) | $3.7x10^{-15}$ | 0.003 (0.008) | 0.73 |
| rs9856387 | 3:38645735 | T/C | 0.30 | 0.031 (0.005) | $1.8x10^{-11}$ | -0.011 (0.008) | 0.15 |
| rs10428132 | 3:38736063 | T/G | 0.41 | -0.062 (0.004) | $5.1x10^{-45}$ | -0.003 (0.007) | 0.72 |
| rs12633819 | 3:66397883 | A/G | 0.66 | -0.043 (0.005) | $2.1x10^{-21}$ | -0.012 (0.007) | 0.11 |
| rs3853159 | 3:69088388 | T/C | 0.48 | -0.025 (0.004) | $3.5x10^{-9}$ | -0.001 (0.007) | 0.86 |
| rs1564756 | 3:69353338 | T/G | 0.27 | -0.031 (0.005) | $2.6x10^{-10}$ | 0.006 (0.008) | 0.46 |
| rs67316928 | 3:89433084 | T/C | 0.61 | 0.043 (0.005) | $5.5x10^{-22}$ | 0.013 (0.007) | 0.081 |
| rs7638138 | 3:111871435 | C/T | 0.33 | -0.055 (0.005) | $2.6x10^{-33}$ | 0.000 (0.008) | 0.99 |
| rs12486285 | 3:122567313 | G/A | 0.74 | 0.034 (0.005) | $1.5x10^{-12}$ | 0.009 (0.008) | 0.25 |
| rs642075 | 3:136215057 | G/A | 0.57 | -0.036 (0.004) | $4.6x10^{-16}$ | -0.013 (0.007) | 0.055 |
| rs6802828 | 3:148997107 | T/C | 0.65 | 0.028 (0.005) | $4.4x10^{-10}$ | 0.011 (0.007) | 0.14 |
| rs7637779 | 3:172257056 | A/G | 0.51 | -0.025 (0.004) | $7.1x10^{-9}$ | 0.008 (0.007) | 0.29 |

| SNP ID | Chomosome: Position | UKB effect/ other allele | UKB effect allele freq | Atrial Fibrillation β (SE) | Atrial Fibrillation P-value | Ischaemic Stroke β (SE) | Ischaemic Stroke P-value |
|---|---|---|---|---|---|---|---|
| rs7612445 | 3:179455191 | G/T | 0.81 | -0.052 (0.006) | $7.4x10^{-21}$ | -0.006 (0.009) | 0.55 |
| rs843376 | 3:184280791 | T/C | 0.41 | 0.025 (0.005) | $2.3x10^{-8}$ | 0.007 (0.007) | 0.34 |
| rs73206619 | 3:195080713 | G/T | 0.76 | -0.048 (0.005) | $3.8x10^{-22}$ | -0.019 (0.009) | 0.027 |
| rs1630029 | 3:196741023 | G/T | 0.39 | 0.025 (0.004) | $1.7x10^{-8}$ | 0.008 (0.007) | 0.27 |
| rs10018140 | 4:1007954 | T/C | 0.29 | 0.031 (0.005) | $2.3x10^{-11}$ | 0.017 (0.008) | 0.045 |
| rs12640611 | 4:10102854 | T/C | 0.30 | -0.038 (0.005) | $2.0x10^{-16}$ | -0.020 (0.008) | 0.012 |
| rs781669 | 4:56953628 | C/T | 0.47 | -0.026 (0.004) | $3.0x10^{-9}$ | -0.010 (0.007) | 0.17 |
| rs12509595 | 4:80261400 | T/C | 0.71 | -0.043 (0.005) | $1.1x10^{-20}$ | -0.028 (0.008) | $1.8x10^{-4}$ |
| rs9307810 | 4:83002657 | A/G | 0.46 | 0.026 (0.004) | $2.2x10^{-9}$ | -0.009 (0.008) | 0.21 |
| rs17309887 | 4:94234354 | C/T | 0.65 | 0.032 (0.005) | $2.2x10^{-12}$ | 0.007 (0.008) | 0.39 |
| rs7660298 | 4:103039611 | T/C | 0.49 | -0.038 (0.004) | $5.1x10^{-19}$ | 0.003 (0.007) | 0.64 |
| rs994978 | 4:110631242 | T/C | 0.31 | 0.148 (0.005) | $2.9x10^{-224}$ | 0.024 (0.008) | $3.0x10^{-3}$ |
| rs2171591 | 4:110798252 | G/A | 0.72 | -0.176 (0.005) | $1.7x10^{-306}$ | -0.030 (0.008) | $9.4x10^{-5}$ |
| rs113832645 | 4:110823850 | G/A | 0.95 | 0.220 (0.011) | $1.8x10^{-87}$ | 0.058 (0.018) | $9.4x10^{-4}$ |
| rs58353956 | 4:111009151 | T/A | 0.98 | 0.081 (0.012) | $1.0x10^{-11}$ | -0.033 (0.028) | 0.24 |
| rs61659434 | 4:148029042 | G/A | 0.94 | -0.098 (0.008) | $1.4x10^{-32}$ | 0.004 (0.014) | 0.74 |
| rs10520260 | 4:173526198 | A/G | 0.68 | 0.039 (0.005) | $8.5x10^{-17}$ | 0.019 (0.008) | 0.017 |
| rs12643389 | 4:173726215 | G/A | 0.92 | 0.083 (0.008) | $4.4x10^{-27}$ | 0.014 (0.015) | 0.35 |
| rs6874933 | 5:188632 | C/T | 0.43 | 0.028 (0.004) | $3.5x10^{-10}$ | -0.004 (0.008) | 0.60 |
| rs71627577 | 5:43125693 | A/G | 0.89 | 0.043 (0.007) | $2.2x10^{-9}$ | -0.006 (0.012) | 0.59 |
| rs34216626 | 5:72640428 | A/T | 0.94 | -0.073 (0.011) | $6.8x10^{-11}$ | -0.016 (0.016) | 0.32 |
| rs4916669 | 5:89097307 | C/T | 0.61 | 0.025 (0.004) | $3.0x10^{-8}$ | 0.003 (0.007) | 0.71 |
| rs2410941 | 5:107115284 | C/A | 0.47 | 0.029 (0.004) | $4.4x10^{-11}$ | 0.002 (0.007) | 0.74 |
| rs2900084 | 5:114421342 | G/A | 0.68 | -0.054 (0.005) | $3.0x10^{-31}$ | -0.019 (0.008) | 0.012 |
| rs285911 | 5:114692761 | C/G | 0.71 | -0.040 (0.006) | $3.7x10^{-12}$ | -0.013 (0.008) | 0.092 |
| rs2287696 | 5:123124637 | G/A | 0.86 | -0.033 (0.006) | $1.2x10^{-8}$ | -0.021 (0.010) | 0.033 |
| rs10055140 | 5:128562825 | A/G | 0.67 | -0.035 (0.005) | $1.8x10^{-11}$ | -0.026 (0.008) | $7.6x10^{-4}$ |
| rs13355516 | 5:138044914 | A/G | 0.83 | -0.118 (0.006) | $1.3x10^{-91}$ | -0.003 (0.009) | 0.72 |
| rs4507481 | 5:138237384 | G/T | 0.75 | -0.033 (0.006) | $1.8x10^{-9}$ | 0.003 (0.009) | 0.76 |
| rs183869 | 5:140151433 | C/T | 0.82 | -0.036 (0.005) | $2.2x10^{-11}$ | -0.009 (0.009) | 0.31 |
| rs11167804 | 5:143122624 | G/T | 0.59 | -0.038 (0.005) | $4.2x10^{-17}$ | -0.022 (0.007) | $2.9x10^{-3}$ |
| rs146462216 | 5:143445440 | G/A | 0.77 | -0.071 (0.005) | $1.2x10^{-41}$ | -0.002 (0.008) | 0.80 |

| SNP ID | Chomosome: Position | UKB effect/ other allele | UKB effect allele freq | Atrial Fibrillation | | Ischaemic Stroke | |
|---|---|---|---|---|---|---|---|
| | | | | β (SE) | P-value | β (SE) | P-value |
| rs62377226 | 5:168965856 | C/T | 0.95 | -0.065 (0.010) | $3.1x10^{-10}$ | -0.023 (0.015) | 0.14 |
| rs6891790 | 5:173243742 | G/T | 0.70 | 0.064 (0.005) | $7.6x10^{-41}$ | 0.018 (0.008) | 0.026 |
| rs876580 | 5:173258036 | T/C | 0.71 | 0.028 (0.005) | $1.4x10^{-8}$ | 0.001 (0.008) | 0.87 |
| rs359477 | 5:173878429 | T/C | 0.56 | 0.038 (0.004) | $5.1x10^{-18}$ | 0.016 (0.007) | 0.023 |
| rs9505044 | 6:7122059 | C/T | 0.94 | -0.053 (0.009) | $1.8x10^{-8}$ | -0.007 (0.015) | 0.64 |
| rs59430691 | 6:16414322 | G/A | 0.86 | 0.104 (0.007) | $4.9x10^{-55}$ | 0.006 (0.011) | 0.59 |
| rs34969716 | 6:18209878 | G/A | 0.69 | -0.070 (0.005) | $2.2x10^{-39}$ | -0.007 (0.008) | 0.43 |
| rs12111199 | 6:22577877 | C/T | 0.72 | -0.030 (0.005) | $4.6x10^{-10}$ | -0.014 (0.008) | 0.079 |
| rs116432905 | 6:31268262 | A/G | 0.93 | 0.045 (0.008) | $2.1x10^{-8}$ | 0.010 (0.013) | 0.44 |
| rs2797964 | 6:34217978 | C/T | 0.09 | 0.057 (0.007) | $1.4x10^{-14}$ | 0.019 (0.014) | 0.18 |
| rs3176326 | 6:36679512 | G/A | 0.80 | 0.068 (0.006) | $1.0x10^{-34}$ | 0.035 (0.009) | $9.4x10^{-5}$ |
| rs77313832 | 6:75295917 | G/A | 0.97 | 0.115 (0.014) | $3.2x10^{-16}$ | 0.020 (0.022) | 0.38 |
| rs9444480 | 6:87169658 | T/C | 0.61 | 0.041 (0.005) | $2.0x10^{-17}$ | -0.001 (0.007) | 0.92 |
| rs9496614 | 6:100165675 | T/C | 0.76 | -0.039 (0.005) | $4.7x10^{-13}$ | -0.007 (0.009) | 0.43 |
| rs72932434 | 6:105131334 | T/C | 0.94 | 0.060 (0.009) | $5.9x10^{-11}$ | 0.046 (0.015) | $2.3x10^{-3}$ |
| rs12195817 | 6:117112474 | T/C | 0.93 | -0.057 (0.009) | $3.2x10^{-11}$ | -0.014 (0.017) | 0.43 |
| rs9481842 | 6:118653635 | T/G | 0.73 | -0.065 (0.005) | $3.7x10^{-39}$ | -0.004 (0.008) | 0.59 |
| rs12529695 | 6:121414130 | A/C | 0.31 | 0.029 (0.005) | $4.1x10^{-10}$ | -0.006 (0.008) | 0.42 |
| rs13191450 | 6:122070990 | A/C | 0.65 | 0.071 (0.005) | $4.3x10^{-56}$ | 0.013 (0.007) | 0.089 |
| rs6941949 | 6:133122523 | C/T | 0.79 | -0.036 (0.005) | $1.2x10^{-11}$ | 0.004 (0.009) | 0.63 |
| rs72980075 | 6:133881068 | T/C | 0.94 | 0.055 (0.009) | $1.9x10^{-9}$ | 0.023 (0.015) | 0.15 |
| rs4896104 | 6:134797951 | C/T | 0.44 | 0.037 (0.004) | $9.5x10^{-17}$ | 0.022 (0.008) | $2.9x10^{-3}$ |
| rs117984853 | 6:149077964 | G/T | 0.91 | -0.113 (0.008) | $1.7x10^{-47}$ | -0.033 (0.013) | $8.7x10^{-3}$ |
| rs10270565 | 7:833467 | C/T | 0.41 | -0.038 (0.004) | $4.8x10^{-18}$ | -0.003 (0.008) | 0.70 |
| rs798502 | 7:2750246 | A/C | 0.71 | 0.028 (0.005) | $1.4x10^{-9}$ | 0.002 (0.008) | 0.76 |
| rs12154315 | 7:14334089 | C/T | 0.74 | -0.049 (0.005) | $1.3x10^{-21}$ | -0.001 (0.009) | 0.93 |
| rs10262140 | 7:27216845 | T/C | 0.06 | -0.045 (0.008) | $1.0x10^{-8}$ | -0.046 (0.014) | $1.2x10^{-3}$ |
| rs4722791 | 7:28372253 | A/C | 0.24 | -0.048 (0.005) | $5.1x10^{-20}$ | -0.004 (0.009) | 0.62 |
| rs11977526 | 7:45968511 | G/A | 0.60 | 0.025 (0.005) | $2.1x10^{-8}$ | 0.024 (0.007) | $1.3x10^{-3}$ |
| rs73137144 | 7:74659260 | A/G | 0.81 | -0.048 (0.007) | $1.8x10^{-12}$ | -0.010 (0.010) | 0.30 |
| rs4729727 | 7:77771272 | T/G | 0.50 | -0.028 (0.004) | $6.3x10^{-11}$ | -0.009 (0.007) | 0.19 |
| rs10232072 | 7:80953280 | T/C | 0.69 | -0.026 (0.005) | $2.0x10^{-8}$ | 0.001 (0.008) | 0.87 |

| SNP ID | Chomosome: Position | UKB effect/ other allele | UKB effect allele freq | Atrial Fibrillation β (SE) | Atrial Fibrillation P-value | Ischaemic Stroke β (SE) | Ischaemic Stroke P-value |
|---|---|---|---|---|---|---|---|
| rs7804293 | 7:92658535 | T/G | 0.74 | 0.048 (0.005) | $7.2 \times 10^{-22}$ | 0.047 (0.008) | $5.0 \times 10^{-9}$ |
| rs4730123 | 7:105972561 | C/T | 0.74 | -0.035 (0.005) | $2.6 \times 10^{-13}$ | 0.006 (0.008) | 0.44 |
| rs4730742 | 7:116497838 | T/G | 0.83 | 0.043 (0.006) | $1.7 \times 10^{-12}$ | 0.010 (0.010) | 0.30 |
| rs3807989 | 7:116546187 | A/G | 0.41 | -0.101 (0.004) | $3.1 \times 10^{-116}$ | -0.018 (0.007) | 0.013 |
| rs55985730 | 7:128776990 | T/G | 0.95 | -0.079 (0.010) | $6.8 \times 10^{-16}$ | -0.029 (0.015) | 0.063 |
| rs7789146 | 7:150964321 | G/A | 0.82 | 0.058 (0.006) | $1.0 \times 10^{-25}$ | 0.018 (0.009) | 0.052 |
| rs10903345 | 8:11941639 | A/G | 0.41 | 0.043 (0.005) | $5.0 \times 10^{-19}$ | -0.003 (0.008) | 0.73 |
| rs7508 | 8:18056461 | G/A | 0.28 | -0.067 (0.005) | $1.1 \times 10^{-44}$ | -0.008 (0.008) | 0.35 |
| rs17060733 | 8:22009422 | T/C | 0.87 | 0.078 (0.007) | $1.3 \times 10^{-33}$ | 0.015 (0.011) | 0.15 |
| rs10503792 | 8:26698083 | G/A | 0.90 | 0.042 (0.008) | $3.9 \times 10^{-8}$ | -0.004 (0.012) | 0.72 |
| rs4737302 | 8:70208042 | A/G | 0.10 | -0.042 (0.007) | $1.4 \times 10^{-10}$ | 0.005 (0.011) | 0.64 |
| rs7005777 | 8:77321364 | G/T | 0.28 | 0.028 (0.005) | $7.9 \times 10^{-9}$ | -0.000 (0.008) | 0.98 |
| rs17181782 | 8:104965515 | T/C | 0.81 | 0.033 (0.005) | $6.8 \times 10^{-10}$ | 0.039 (0.009) | $6.7 \times 10^{-6}$ |
| rs12677136 | 8:117866891 | T/C | 0.83 | -0.039 (0.006) | $6.5 \times 10^{-12}$ | -0.003 (0.010) | 0.73 |
| rs78332318 | 8:123533862 | C/T | 0.94 | -0.120 (0.010) | $1.0 \times 10^{-36}$ | -0.021 (0.015) | 0.15 |
| rs7460121 | 8:134800173 | G/A | 0.90 | -0.064 (0.008) | $9.1 \times 10^{-17}$ | 0.001 (0.013) | 0.93 |
| rs4961234 | 8:140830782 | T/C | 0.55 | -0.039 (0.004) | $2.2 \times 10^{-19}$ | -0.028 (0.007) | $8.2 \times 10^{-5}$ |
| rs4237169 | 9:8895340 | A/G | 0.43 | -0.032 (0.005) | $4.5 \times 10^{-13}$ | 0.001 (0.007) | 0.89 |
| rs4961747 | 9:16744271 | G/A | 0.79 | -0.030 (0.005) | $3.5 \times 10^{-8}$ | 0.003 (0.009) | 0.78 |
| rs1339552 | 9:16848792 | C/T | 0.60 | -0.027 (0.004) | $8.1 \times 10^{-10}$ | -0.016 (0.007) | 0.024 |
| rs398471 | 9:20215707 | G/A | 0.42 | -0.036 (0.005) | $7.8 \times 10^{-15}$ | -0.005 (0.007) | 0.47 |
| rs7864171 | 9:94960984 | G/A | 0.58 | -0.075 (0.004) | $8.1 \times 10^{-66}$ | -0.006 (0.007) | 0.41 |
| rs12375999 | 9:96325514 | C/T | 0.84 | -0.033 (0.006) | $3.8 \times 10^{-8}$ | 0.009 (0.010) | 0.34 |
| rs7874948 | 9:98864407 | A/C | 0.27 | -0.029 (0.005) | $1.6 \times 10^{-8}$ | -0.018 (0.008) | 0.031 |
| rs3780197 | 9:124413442 | T/C | 0.56 | 0.026 (0.004) | $1.6 \times 10^{-9}$ | 0.013 (0.007) | 0.068 |
| rs4842131 | 9:136200833 | T/C | 0.42 | -0.034 (0.004) | $1.2 \times 10^{-14}$ | -0.005 (0.008) | 0.48 |
| rs2265099 | 10:22207976 | C/A | 0.74 | -0.029 (0.005) | $3.6 \times 10^{-9}$ | 0.008 (0.008) | 0.34 |
| rs2484726 | 10:32758334 | T/C | 0.11 | -0.040 (0.007) | $1.1 \times 10^{-9}$ | -0.001 (0.012) | 0.93 |
| rs12253763 | 10:49221982 | C/T | 0.91 | 0.058 (0.007) | $9.1 \times 10^{-16}$ | 0.000 (0.011) | 0.97 |
| rs4838500 | 10:49334177 | T/C | 0.17 | -0.033 (0.006) | $1.0 \times 10^{-8}$ | -0.016 (0.010) | 0.11 |
| rs7898861 | 10:63559918 | T/C | 0.52 | 0.051 (0.004) | $2.2 \times 10^{-32}$ | -0.001 (0.007) | 0.91 |
| rs10997728 | 10:67570594 | T/C | 0.88 | -0.055 (0.006) | $9.1 \times 10^{-19}$ | 0.012 (0.010) | 0.23 |

| SNP ID | Chomosome: Position | UKB effect/ other allele | UKB effect allele freq | Atrial Fibrillation β (SE) | Atrial Fibrillation P-value | Ischaemic Stroke β (SE) | Ischaemic Stroke P-value |
|---|---|---|---|---|---|---|---|
| rs60632610 | 10:73655919 | C/T | 0.86 | 0.116 (0.006) | $1.0x10^{-82}$ | 0.012 (0.010) | 0.23 |
| rs6480770 | 10:75098799 | A/G | 0.60 | 0.026 (0.005) | $3.8x10^{-9}$ | -0.005 (0.007) | 0.52 |
| rs11001667 | 10:76175587 | A/G | 0.83 | -0.053 (0.006) | $5.5x10^{-22}$ | 0.001 (0.010) | 0.95 |
| rs3802662 | 10:86685984 | C/T | 0.30 | -0.032 (0.005) | $4.3x10^{-12}$ | 0.000 (0.008) | 0.96 |
| rs2138042 | 10:89510130 | G/A | 0.83 | -0.033 (0.006) | $2.6x10^{-9}$ | -0.002 (0.009) | 0.83 |
| rs1044258 | 10:101845957 | T/C | 0.66 | 0.034 (0.005) | $6.5x10^{-14}$ | -0.001 (0.008) | 0.88 |
| rs71471272 | 10:103245744 | G/A | 0.93 | 0.051 (0.009) | $2.0x10^{-8}$ | -0.029 (0.015) | 0.043 |
| rs11598047 | 10:103582915 | A/G | 0.85 | -0.163 (0.006) | $1.9x10^{-174}$ | -0.031 (0.010) | $1.6x10^{-3}$ |
| rs80056983 | 10:103750144 | C/T | 0.88 | -0.122 (0.006) | $2.5x10^{-85}$ | -0.037 (0.011) | $6.8x10^{-4}$ |
| rs3121481 | 10:113646548 | A/G | 0.37 | -0.030 (0.005) | $1.0x10^{-9}$ | -0.004 (0.007) | 0.57 |
| rs12571587 | 10:114655839 | T/C | 0.49 | 0.030 (0.004) | $2.6x10^{-12}$ | 0.014 (0.007) | 0.040 |
| rs10787851 | 10:118665783 | T/C | 0.67 | 0.026 (0.005) | $4.5x10^{-9}$ | 0.005 (0.008) | 0.48 |
| rs2981582 | 10:121592803 | A/G | 0.39 | -0.031 (0.004) | $1.1x10^{-12}$ | -0.002 (0.007) | 0.84 |
| rs4980389 | 11:1871355 | G/A | 0.62 | -0.026 (0.004) | $2.7x10^{-9}$ | -0.028 (0.007) | $1.8x10^{-4}$ |
| rs10835185 | 11:3841113 | G/T | 0.73 | 0.035 (0.006) | $8.9x10^{-10}$ | 0.004 (0.009) | 0.68 |
| rs10832172 | 11:14050863 | T/C | 0.52 | -0.035 (0.004) | $1.1x10^{-16}$ | -0.002 (0.007) | 0.80 |
| rs2625322 | 11:19988805 | G/A | 0.77 | -0.070 (0.005) | $6.6x10^{-44}$ | -0.020 (0.009) | 0.018 |
| rs11031120 | 11:30515959 | C/T | 0.74 | -0.030 (0.005) | $1.1x10^{-10}$ | 0.001 (0.008) | 0.93 |
| rs11038350 | 11:45200412 | C/G | 0.71 | -0.029 (0.005) | $3.2x10^{-8}$ | -0.009 (0.008) | 0.23 |
| rs10791902 | 11:67325889 | C/T | 0.59 | 0.025 (0.005) | $2.2x10^{-8}$ | 0.001 (0.008) | 0.87 |
| rs12798693 | 11:69278268 | A/G | 0.51 | -0.026 (0.004) | $1.6x10^{-9}$ | -0.010 (0.007) | 0.16 |
| rs3015960 | 11:69498730 | T/C | 0.68 | -0.029 (0.005) | $2.3x10^{-9}$ | 0.016 (0.008) | 0.053 |
| rs559019 | 11:95362500 | G/A | 0.28 | 0.033 (0.005) | $1.4x10^{-12}$ | 0.003 (0.008) | 0.74 |
| rs60016141 | 11:113792288 | G/A | 0.92 | -0.048 (0.007) | $2.7x10^{-11}$ | -0.005 (0.012) | 0.70 |
| rs6589894 | 11:121765352 | C/T | 0.26 | 0.032 (0.005) | $9.5x10^{-11}$ | 0.030 (0.008) | $2.9x10^{-4}$ |
| rs76097649 | 11:128894675 | G/A | 0.91 | -0.107 (0.008) | $5.2x10^{-43}$ | -0.005 (0.013) | 0.71 |
| rs12813456 | 12:3972130 | G/A | 0.80 | 0.042 (0.005) | $5.9x10^{-15}$ | 0.006 (0.009) | 0.53 |
| rs76895963 | 12:4275678 | T/G | 0.98 | -0.178 (0.020) | $2.1x10^{-18}$ | 0.079 (0.031) | $9.7x10^{-3}$ |
| rs10845620 | 12:12733093 | G/A | 0.88 | -0.050 (0.007) | $1.5x10^{-13}$ | 0.005 (0.012) | 0.64 |
| rs10842383 | 12:24619033 | C/T | 0.85 | 0.099 (0.006) | $1.6x10^{-57}$ | 0.012 (0.010) | 0.23 |
| rs58747679 | 12:26195371 | T/C | 0.71 | 0.045 (0.005) | $1.1x10^{-21}$ | 0.002 (0.008) | 0.81 |
| rs28698947 | 12:26569837 | A/G | 0.80 | -0.042 (0.005) | $1.5x10^{-14}$ | 0.000 (0.009) | 0.97 |

| SNP ID | Chomosome: Position | UKB effect/ other allele | UKB effect allele freq | Atrial Fibrillation β (SE) | Atrial Fibrillation P-value | Ischaemic Stroke β (SE) | Ischaemic Stroke P-value |
|---|---|---|---|---|---|---|---|
| rs1026372 | 12:32815256 | T/C | 0.23 | 0.034 (0.005) | $2.9x10^{-11}$ | 0.003 (0.009) | 0.70 |
| rs75704921 | 12:45906317 | C/T | 0.97 | -0.091 (0.014) | $4.7x10^{-11}$ | -0.055 (0.024) | 0.020 |
| rs12379 | 12:51049161 | G/A | 0.63 | -0.025 (0.005) | $1.8x10^{-8}$ | 0.002 (0.007) | 0.78 |
| rs7978685 | 12:56709370 | T/C | 0.27 | 0.055 (0.005) | $4.4x10^{-30}$ | 0.003 (0.008) | 0.73 |
| rs42874 | 12:69620934 | T/C | 0.59 | 0.043 (0.004) | $2.6x10^{-22}$ | -0.003 (0.007) | 0.69 |
| rs111233078 | 12:69681938 | G/A | 0.61 | 0.027 (0.004) | $7.2x10^{-10}$ | 0.001 (0.007) | 0.85 |
| rs1565765 | 12:75845075 | C/T | 0.50 | 0.040 (0.004) | $7.1x10^{-20}$ | 0.014 (0.007) | 0.045 |
| rs2681485 | 12:89631845 | G/A | 0.41 | -0.025 (0.004) | $1.1x10^{-8}$ | -0.019 (0.007) | 0.011 |
| rs2466551 | 12:104119140 | T/G | 0.85 | 0.045 (0.006) | $2.5x10^{-14}$ | 0.013 (0.010) | 0.18 |
| rs4964188 | 12:106706939 | T/C | 0.50 | 0.024 (0.004) | $1.5x10^{-8}$ | 0.005 (0.007) | 0.46 |
| rs4766552 | 12:111193961 | G/A | 0.94 | 0.061 (0.011) | $3.9x10^{-8}$ | 0.004 (0.014) | 0.77 |
| rs883079 | 12:114355435 | C/T | 0.27 | -0.101 (0.005) | $1.0x10^{-102}$ | -0.015 (0.008) | 0.053 |
| rs7955305 | 12:114433886 | G/A | 0.91 | -0.065 (0.008) | $7.9x10^{-18}$ | -0.034 (0.012) | $5.7x10^{-3}$ |
| rs1061657 | 12:114670331 | T/C | 0.75 | -0.043 (0.006) | $3.7x10^{-15}$ | 0.010 (0.009) | 0.23 |
| rs3768 | 12:124015292 | C/T | 0.81 | 0.046 (0.006) | $3.2x10^{-15}$ | 0.024 (0.009) | $8.8x10^{-3}$ |
| rs7296889 | 12:132478070 | T/C | 0.49 | -0.030 (0.004) | $1.8x10^{-11}$ | -0.009 (0.008) | 0.23 |
| rs2798280 | 13:21538714 | T/C | 0.60 | -0.033 (0.005) | $6.2x10^{-12}$ | 0.010 (0.007) | 0.16 |
| rs9506925 | 13:22794804 | C/T | 0.73 | -0.054 (0.005) | $3.0x10^{-26}$ | -0.010 (0.008) | 0.21 |
| rs1773133 | 13:46647085 | G/C | 0.32 | -0.035 (0.005) | $5.1x10^{-12}$ | -0.017 (0.008) | 0.030 |
| rs9573330 | 13:73944073 | G/A | 0.63 | -0.027 (0.005) | $1.4x10^{-9}$ | -0.013 (0.007) | 0.078 |
| rs9557850 | 13:102290788 | C/T | 0.71 | 0.031 (0.005) | $3.5x10^{-10}$ | -0.005 (0.008) | 0.56 |
| rs35569628 | 13:113218398 | T/C | 0.77 | 0.028 (0.005) | $3.5x10^{-8}$ | 0.006 (0.008) | 0.44 |
| rs41306688 | 13:113424243 | A/C | 0.97 | -0.069 (0.012) | $1.9x10^{-8}$ | -0.041 (0.022) | 0.065 |
| rs28631169 | 14:23418974 | C/T | 0.80 | -0.057 (0.006) | $8.5x10^{-24}$ | -0.002 (0.009) | 0.83 |
| rs7493664 | 14:32467172 | A/C | 0.31 | -0.034 (0.005) | $2.1x10^{-12}$ | -0.005 (0.009) | 0.57 |
| rs7140396 | 14:32514611 | G/A | 0.73 | -0.066 (0.005) | $1.6x10^{-44}$ | -0.008 (0.008) | 0.36 |
| rs8005490 | 14:34715335 | T/C | 0.41 | 0.045 (0.004) | $2.2x10^{-24}$ | 0.013 (0.007) | 0.063 |
| rs2738413 | 14:64213242 | A/G | 0.48 | 0.068 (0.004) | $7.8x10^{-57}$ | 0.012 (0.007) | 0.090 |
| rs4903064 | 14:72812712 | T/C | 0.76 | 0.043 (0.005) | $1.1x10^{-16}$ | 0.004 (0.009) | 0.65 |
| rs10873298 | 14:76960182 | C/T | 0.39 | 0.044 (0.005) | $4.0x10^{-19}$ | 0.017 (0.008) | 0.032 |
| rs17796831 | 14:85388136 | C/T | 0.73 | -0.028 (0.005) | $1.4x10^{-8}$ | -0.009 (0.008) | 0.31 |
| rs179152 | 14:95515273 | C/T | 0.74 | -0.031 (0.005) | $9.3x10^{-10}$ | -0.009 (0.008) | 0.30 |

| SNP ID | Chomosome: Position | UKB effect/ other allele | UKB effect allele freq | Atrial Fibrillation β (SE) | Atrial Fibrillation P-value | Ischaemic Stroke β (SE) | Ischaemic Stroke P-value |
|---|---|---|---|---|---|---|---|
| rs117435437 | 15:57484473 | T/A | 0.97 | -0.086 (0.015) | $2.3x10^{-8}$ | 0.012 (0.024) | 0.62 |
| rs62011291 | 15:63507814 | A/G | 0.79 | -0.030 (0.005) | $1.2x10^{-8}$ | -0.003 (0.008) | 0.70 |
| rs6494611 | 15:66851514 | G/A | 0.64 | -0.028 (0.005) | $3.5x10^{-10}$ | -0.019 (0.007) | 0.011 |
| rs74022964 | 15:73384923 | C/T | 0.84 | -0.111 (0.006) | $1.0x10^{-77}$ | -0.018 (0.010) | 0.069 |
| rs8028676 | 15:80375025 | C/A | 0.60 | -0.033 (0.004) | $2.3x10^{-14}$ | -0.023 (0.007) | $1.1x10^{-3}$ |
| rs7166287 | 15:98729846 | C/T | 0.37 | 0.037 (0.004) | $7.4x10^{-17}$ | 0.002 (0.007) | 0.78 |
| rs7205337 | 16:730339 | A/G | 0.78 | -0.030 (0.005) | $6.6x10^{-9}$ | 0.000 (0.009) | 0.99 |
| rs140185678 | 16:1953015 | G/A | 0.97 | -0.184 (0.015) | $5.4x10^{-37}$ | 0.046 (0.023) | 0.046 |
| rs36232 | 16:2149787 | T/G | 0.19 | -0.041 (0.006) | $7.2x10^{-12}$ | -0.003 (0.010) | 0.76 |
| rs9284324 | 16:15808858 | G/A | 0.68 | 0.033 (0.005) | $3.2x10^{-12}$ | -0.012 (0.008) | 0.13 |
| rs62037428 | 16:28996758 | C/T | 0.65 | 0.029 (0.005) | $1.0x10^{-9}$ | 0.023 (0.009) | $9.0x10^{-3}$ |
| rs1558902 | 16:53769662 | T/A | 0.61 | -0.039 (0.005) | $9.5x10^{-16}$ | -0.012 (0.009) | 0.17 |
| rs117285855 | 16:72262694 | T/C | 0.98 | -0.097 (0.018) | $4.7x10^{-8}$ | -0.084 (0.032) | $8.7x10^{-3}$ |
| rs4404097 | 16:72998133 | G/A | 0.85 | -0.177 (0.006) | $6.2x10^{-213}$ | -0.034 (0.010) | $4.5x10^{-4}$ |
| rs11075959 | 16:73001146 | A/G | 0.02 | -0.126 (0.014) | $1.9x10^{-18}$ | 0.032 (0.028) | 0.25 |
| rs62055075 | 16:73051262 | G/A | 0.96 | 0.069 (0.010) | $2.6x10^{-11}$ | 0.023 (0.021) | 0.29 |
| rs931458 | 16:86372699 | A/C | 0.46 | 0.027 (0.004) | $2.7x10^{-10}$ | 0.007 (0.007) | 0.33 |
| rs74776107 | 17:1370609 | T/G | 0.92 | 0.063 (0.007) | $2.3x10^{-17}$ | -0.006 (0.013) | 0.67 |
| rs2209072 | 17:2302425 | A/C | 0.38 | 0.033 (0.004) | $8.1x10^{-14}$ | 0.005 (0.007) | 0.48 |
| rs57985740 | 17:7510306 | A/G | 0.79 | -0.036 (0.006) | $1.4x10^{-10}$ | -0.021 (0.009) | 0.017 |
| rs55941572 | 17:12709614 | T/C | 0.89 | 0.069 (0.007) | $1.1x10^{-21}$ | 0.002 (0.012) | 0.84 |
| rs9635726 | 17:39863888 | C/T | 0.80 | -0.037 (0.005) | $1.8x10^{-12}$ | 0.002 (0.009) | 0.83 |
| rs17608766 | 17:46935905 | T/C | 0.86 | -0.064 (0.007) | $7.8x10^{-23}$ | -0.051 (0.010) | $2.8x10^{-7}$ |
| rs9906486 | 17:66194802 | G/T | 0.95 | -0.070 (0.009) | $5.8x10^{-15}$ | -0.010 (0.016) | 0.54 |
| rs9912702 | 17:70452588 | C/A | 0.48 | -0.032 (0.004) | $2.2x10^{-13}$ | -0.017 (0.007) | 0.013 |
| rs56085832 | 18:2823279 | G/A | 0.89 | -0.041 (0.007) | $9.8x10^{-9}$ | -0.020 (0.012) | 0.086 |
| rs2230234 | 18:31524751 | A/G | 0.92 | -0.066 (0.008) | $1.4x10^{-15}$ | -0.008 (0.012) | 0.51 |
| rs878767 | 18:34841675 | A/G | 0.74 | -0.033 (0.005) | $2.9x10^{-11}$ | 0.003 (0.008) | 0.68 |
| rs9953366 | 18:48947822 | T/C | 0.32 | -0.041 (0.005) | $9.3x10^{-19}$ | -0.027 (0.008) | $5.9x10^{-4}$ |
| rs4799053 | 18:79398019 | A/G | 0.20 | -0.034 (0.006) | $2.4x10^{-9}$ | 0.001 (0.010) | 0.90 |
| rs740404 | 19:2232050 | G/A | 0.88 | -0.042 (0.007) | $8.3x10^{-10}$ | -0.007 (0.012) | 0.58 |
| rs2620797 | 19:5087118 | C/T | 0.94 | -0.055 (0.009) | $6.8x10^{-10}$ | -0.011 (0.014) | 0.44 |

| SNP ID | Chomosome: Position | UKB effect/ other allele | UKB effect allele freq | Atrial Fibrillation β (SE) | Atrial Fibrillation P-value | Ischaemic Stroke β (SE) | Ischaemic Stroke P-value |
|---|---|---|---|---|---|---|---|
| rs62109758 | 19:36100514 | A/G | 0.90 | 0.043 (0.007) | $8.3x10^{-9}$ | 0.022 (0.012) | 0.058 |
| rs2911003 | 19:47664678 | T/C | 0.32 | -0.037 (0.005) | $2.5x10^{-15}$ | -0.003 (0.008) | 0.76 |
| rs4801459 | 19:57303321 | C/T | 0.78 | -0.031 (0.005) | $3.9x10^{-9}$ | -0.011 (0.009) | 0.20 |
| rs2145274 | 20:6591367 | A/C | 0.92 | 0.076 (0.009) | $8.5x10^{-18}$ | 0.026 (0.014) | 0.055 |
| rs6082361 | 20:21265982 | G/A | 0.35 | 0.029 (0.005) | $2.5x10^{-10}$ | 0.014 (0.008) | 0.073 |
| rs6092091 | 20:38208389 | T/C | 0.77 | 0.029 (0.005) | $1.5x10^{-8}$ | 0.002 (0.008) | 0.82 |
| rs2092518 | 20:44186048 | G/T | 0.56 | -0.032 (0.004) | $2.6x10^{-13}$ | -0.022 (0.007) | $1.9x10^{-3}$ |
| rs57430487 | 20:49351083 | T/C | 0.92 | -0.057 (0.010) | $7.4x10^{-9}$ | -0.019 (0.014) | 0.17 |
| rs6122011 | 20:62559739 | G/T | 0.59 | 0.037 (0.005) | $7.2x10^{-17}$ | 0.008 (0.008) | 0.29 |
| rs2834618 | 21:34746814 | T/G | 0.90 | 0.103 (0.007) | $1.3x10^{-45}$ | 0.014 (0.012) | 0.24 |
| rs2836961 | 21:39255094 | A/C | 0.63 | 0.037 (0.005) | $1.6x10^{-16}$ | 0.012 (0.007) | 0.10 |
| rs75292526 | 21:44345083 | C/T | 0.94 | -0.053 (0.010) | $3.0x10^{-8}$ | -0.011 (0.016) | 0.51 |
| rs464901 | 22:18114735 | T/C | 0.65 | 0.044 (0.005) | $2.5x10^{-17}$ | 0.013 (0.008) | 0.098 |
| rs133885 | 22:25763322 | G/A | 0.54 | -0.036 (0.004) | $3.0x10^{-16}$ | -0.012 (0.009) | 0.20 |
| rs1004243 | 22:31101893 | C/T | 0.59 | 0.027 (0.005) | $4.8x10^{-9}$ | 0.008 (0.007) | 0.28 |
| rs7292074 | 22:38870446 | C/A | 0.81 | -0.035 (0.005) | $8.7x10^{-11}$ | -0.003 (0.009) | 0.74 |

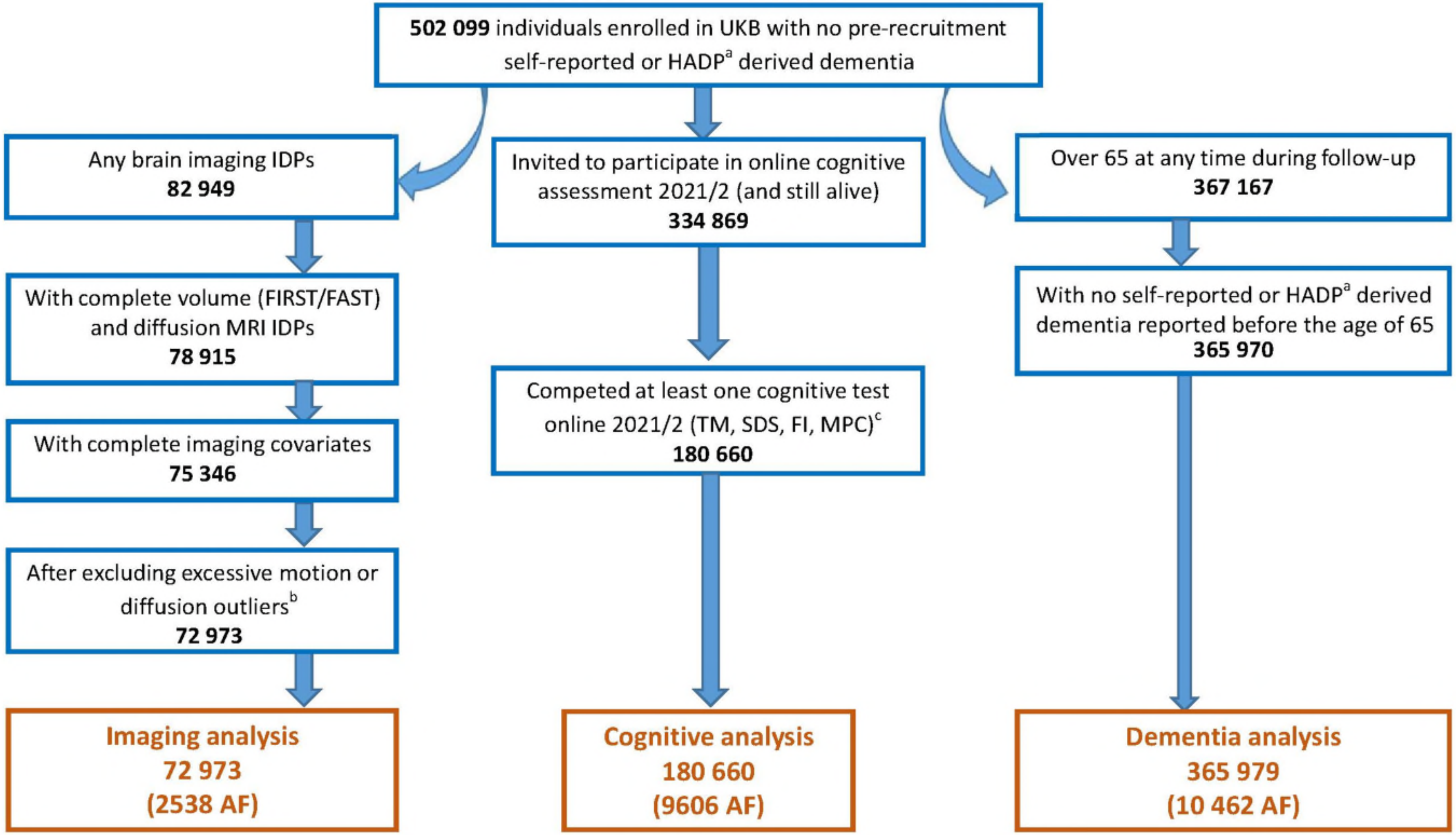


**eFigure 1: Study design showing the number of individuals in each analysis and the associated exclusions.**
[a]Electronic hospital admissions and procedure records [b]Limits calculated using all complete cases – resting function mean head motion >3 x standard deviation or number of diffusion outliers > 3 x standard deviation. [c]Trail making, symbol digit substitution, fluid intelligence and matrix pattern completion.

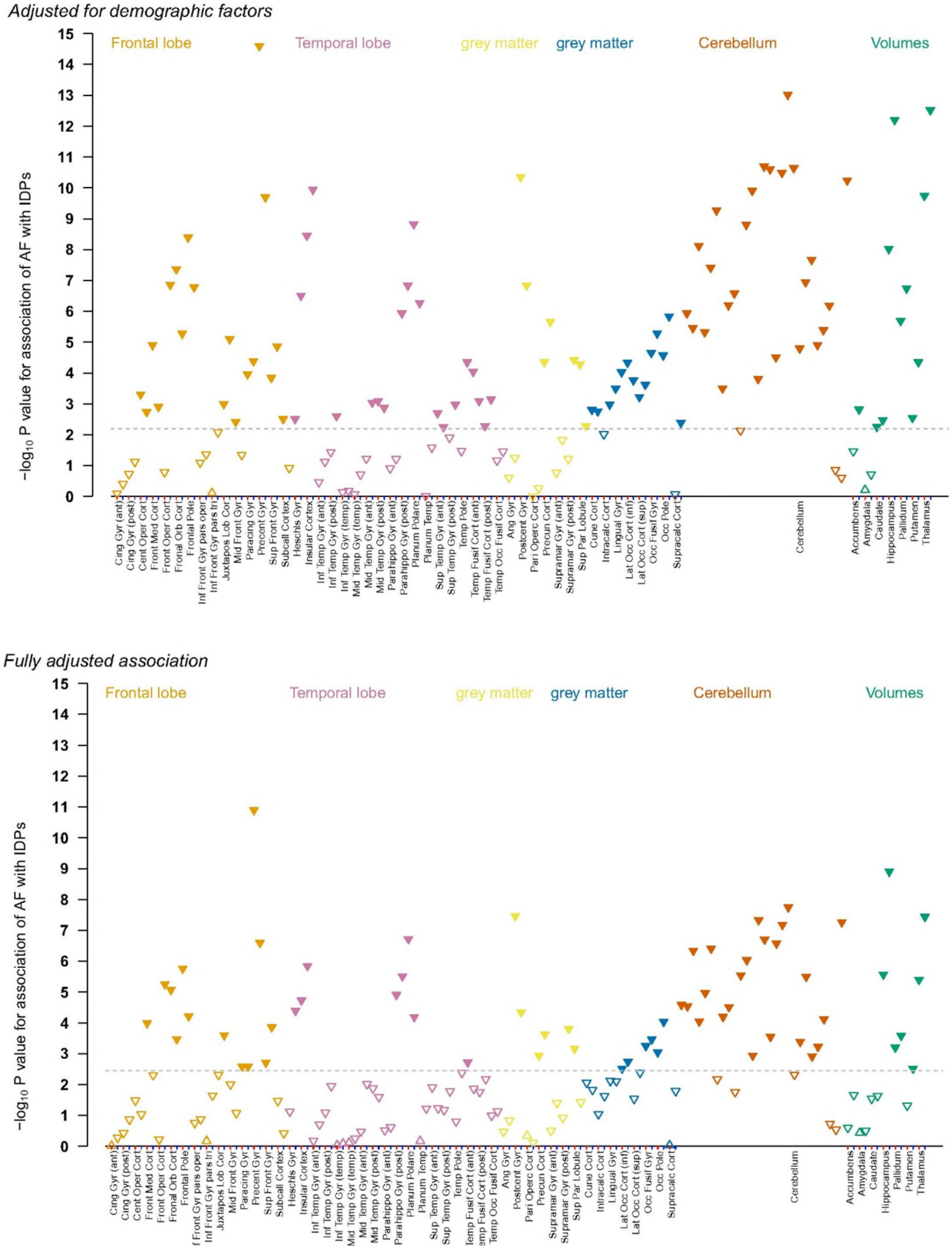


**eFigure 2: The association between AF (n=2538) and regional brain volume IDPs in 72 073 individuals (a) adjusted for demographic factors and (b) fully adjusted.**
All analyses are adjusted for age at imaging, sex, education and deprivation plus technical imaging confounders (supplementary table 3). Fully adjusted analyses are also adjusted for CVD risk factors, other pre-imaging cardiovascular comobidities including haemorrhagic and ischaemic stroke. Downward pointing arrows indicate that the estimate is negative, upwards pointing arrows indicate a positive relationship. Raw p-values are shown. Filled arrows are above FDR significance (Benjami-Yekutieli-adjusted p-value<0.05, corresponding to raw p-values shown by the dashed lines).

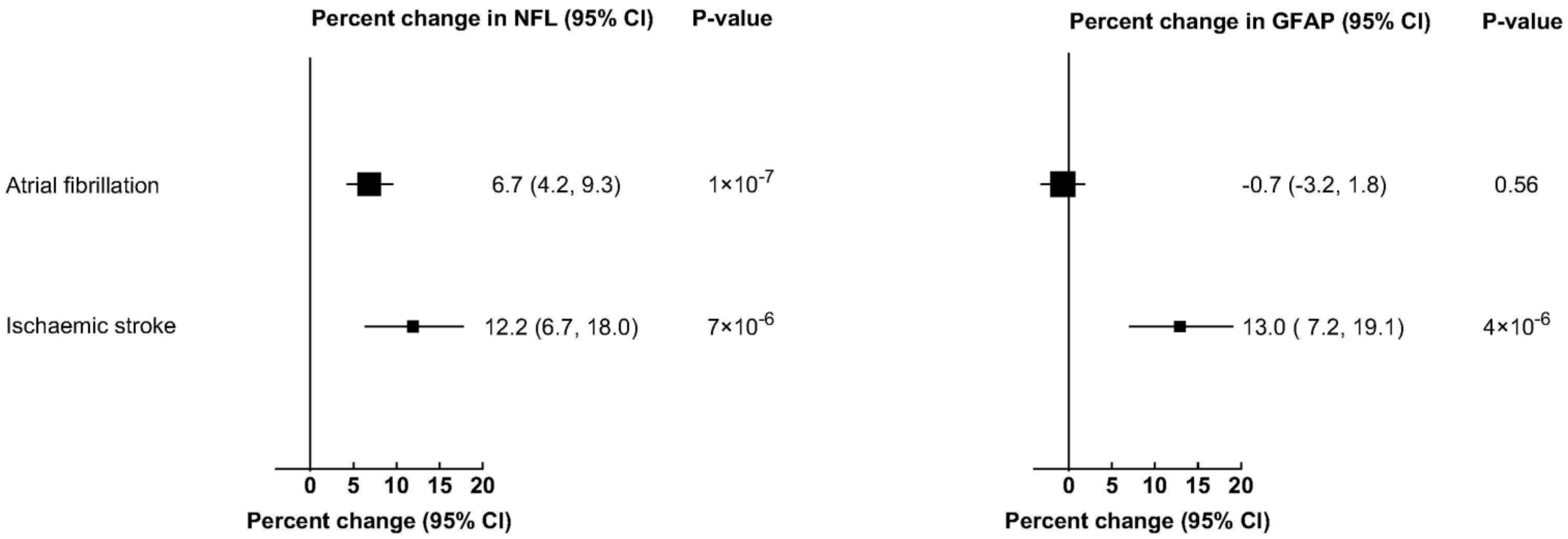


**eFigure 3: Fully adjusted association of (a) recruitment NFL or (b) recruitment GFAP with prevalent AF and ischaemic stroke**
